# Reducing misclassification bias in electronic health record-based GWAS of psychiatric traits using the SuperControl framework

**DOI:** 10.64898/2025.12.14.25342213

**Authors:** Lisa Eick, FinnGen, Andrea Ganna, Zhiyu Yang

**Affiliations:** Institute for Molecular Medicine Finland, Helsinki Institute of Life Science, University of Helsinki, Helsinki, Finland

## Abstract

Electronic health records (EHRs) have enabled large-scale genetic studies of psychiatric disorders, but their diagnostic imprecision introduces substantial misclassification bias. This challenge is particularly pronounced for psychiatric traits, which lack objective biomarkers, exhibit high comorbidity, and are often underdiagnosed due to stigma and help-seeking barriers. We first used simulations to examine how misclassification bias interacts with the classic “super-normal” control design. The results showed that, under moderate to high misclassification, especially for more prevalent traits, excluding individuals with psychiatric comorbidities from the controls can recover power and reduce false positives. Guided by these insights, we constructed a refined SuperControl phenotype in FinnGen for ten psychiatric disorders. Despite reduced sample size, this approach increased genome-wide locus discovery and enhanced biological specificity, with stronger CNS heritability enrichment and improved cross-biobank polygenic prediction, while modestly increasing genetic correlations between traits. We further developed PRISMA, a machine-learning approach that complements the SuperControl phenotype by leveraging excluded individuals through imputation of disease liability. PRISMA further increased locus discovery, revealing significant associations for previously underpowered traits, though with greater pleiotropy and reduced tissue specificity. Together, these findings demonstrate that misclassification in EHR-derived psychiatric phenotypes can meaningfully suppress genetic signal, and that structured control refinement can mitigate this bias. Our results suggest that stricter cohort selection, supplemented by predictive imputation when appropriate, offers a scalable strategy to enhance discovery and generalizability in EHR-based psychiatric genetics.

## Introduction

Large-scale biobanks linked to electronic health records (EHR) have enabled genome-wide association studies (GWAS) across hundreds of diseases. The use of EHR to define disease outcomes in GWAS introduces several challenges. EHR systems are shaped by country-specific healthcare practices, diagnostic coding standards, and data collection procedures, which can limit cross-cohort comparability. Moreover, EHR-derived disease definitions often rely on binary diagnostic codes that may overlook subthreshold symptoms, obscure clinical nuance, or include misclassified individuals due to incomplete medical histories, diagnostic bias, or societal stigma^1,2^. These limitations are particularly acute for psychiatric traits, which lack definitive biomarkers and are prone to underdiagnosis or misdiagnosis. As a result, many EHR-based psychiatric traits are considered *shallow*, defined by large cohort sizes but low specificity^3–5^. Although it is often assumed that large sample sizes will compensate for this misclassification, prior work has shown that even modest levels of misclassification can attenuate genetic signals and bias downstream analyses^6^.

A growing body of research has sought to counteract these limitations by creating *deep phenotypes*, more accurate, fine-grained trait definitions that better capture true disease status. Many successful efforts have relied on supplemental data sources, such as questionnaires, family history, or clinical interviews^2,7,8^, but these are typically available only for a subset of participants, reducing effective sample size. Recent studies have addressed this by imputing deep phenotypes for the remaining participants using predictive models^2,7–10^. While these approaches can improve specificity, their reliance on additional data limits applicability across EHR-based biobanks.

An alternative strategy, rooted in early psychiatric epidemiology, is the use of *super-normal controls*: control groups screened to exclude not only the focal disorder but also any other psychiatric condition^11,12^. This design was proposed to increase statistical power in genetic studies of psychiatric traits, which have historically been underpowered. While effective in enhancing signal, it has been criticized for introducing bias by producing non-representative control samples^13–15^. To date, however, the relationships between misclassification bias in EHR-derived phenotypes and the super-normal control design has not been systematically evaluated.

We address this gap by first using simulations to test whether super-normal control definitions can mitigate the effects of EHR-related misclassification. We then apply this approach to nine major psychiatric trait and a composite trait of any mental health disorder in FinnGen (N=508,365), systematically assessing its impact on genome-wide discovery, replication, cross-biobank transferability, and tissue-specific heritability. Finally, to gain statistical power while retaining the benefits of the SuperControl approach, we develop a machine learning framework, PRISMA (*PRedicted excluded individuals and SuperControl cohort integrated via MTAG Analysis*), that imputes the phenotype of individuals from the SuperControl approach. Together, our work provides a practical and scalable strategy to improve genetic studies of underrepresented and misclassified traits in EHR data, with particular relevance for psychiatric traits.

## Results

### Simulations show that the SuperControl definition can mitigate misclassification Bias in GWAS

To evaluate the impact of misclassification in GWAS and the performance of the SuperControl design, formerly known as the *super-normal control* design in psychiatric epidemiology, we conducted simulations under varying levels of diagnostic error (Fig. 1). This allowed us to address two key questions: do misclassification bias and SuperControl bias act independently or interact, and can the SuperControl strategy offset misclassification without introducing additional distortion?

**Fig. 1.**
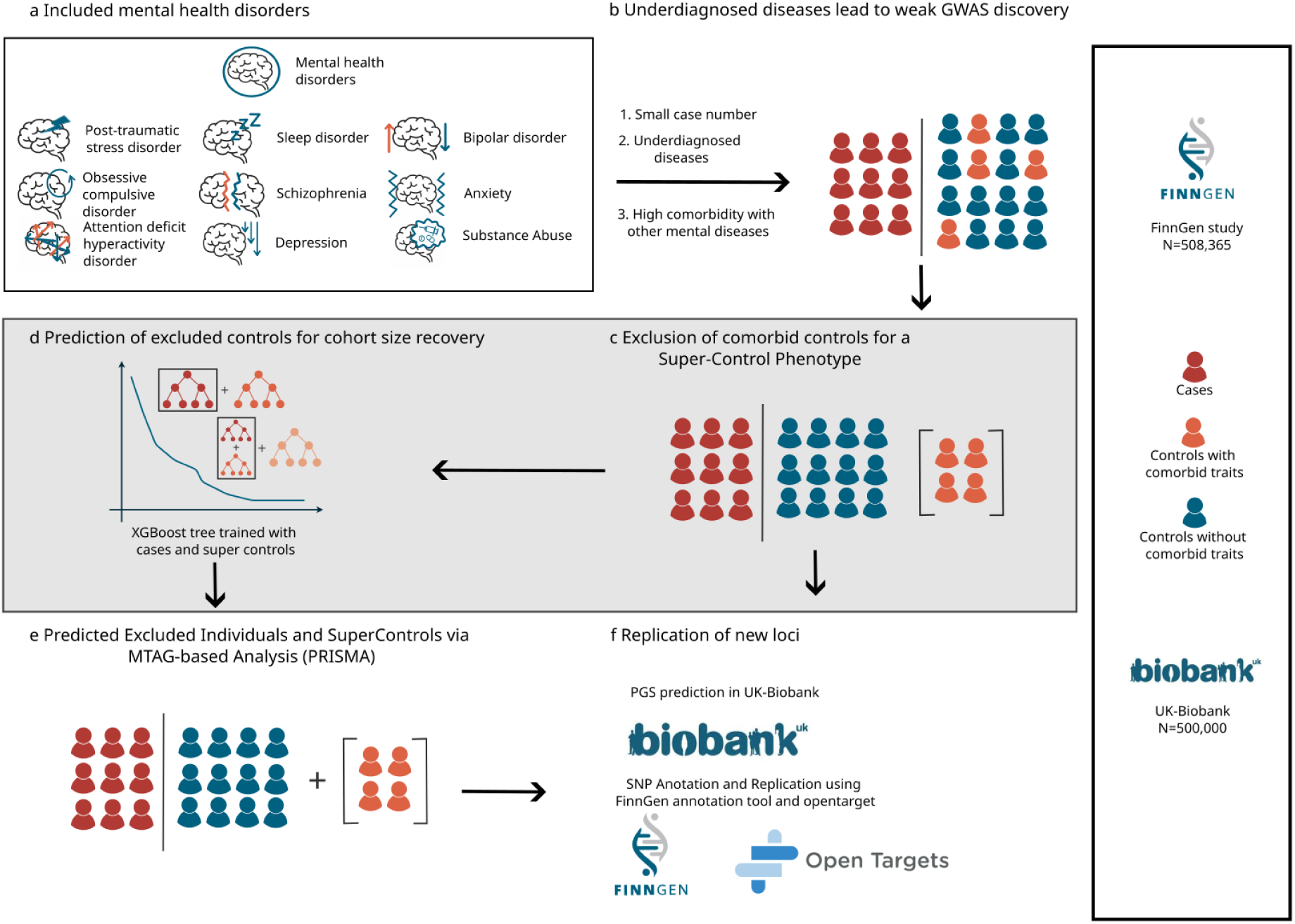
Study overview and phenotype refinement workflow. **a**, Overview of ten tested psychiatric traits with diagnostic uncertainty or low prevalence: PTSD, sleep disorders, bipolar disorder, anxiety, substance abuse, depression, schizophrenia, OCD, ADHD, and general mental illness. **b**, Traditional GWAS suffer from reduced power due to underdiagnosis and comorbidities, which blur case-control distinctions. **c**, Excluding individuals with high psychiatric comorbidity yields a “super control” phenotype with cleaner case-control contrast. **d**, An XGBoost model predicts disease status in excluded individuals based on the super-control phenotype. **e**, Predicted cases are combined with the super-control GWAS using MTAG to create the Imputed Case-Control Phenotype (PRISMA), restoring full cohort power. **f**, Replication and biological relevance of new loci are assessed using polygenic scoring in UK Biobank and functional annotation tools like OpenTargets. ADHD, attention-deficit/hyperactivity disorder; GWAS, genome-wide association study; MTAG, multi-trait analysis of GWAS; OCD, obsessive-compulsive disorder; PTSD, post-traumatic stress disorder;

We simulated two binary traits: trait A, representing the disorder of interest, and trait B, a comorbid condition used to define SuperControls. We assumed trait A and B share some causal genetic signals, whereas each of them also have their trait specific genetics. Standard controls included all individuals unaffected by trait A, whereas SuperControls excluded individuals affected by either trait A or trait B. Each design was tested with and without misclassification, and the performance was evaluated by power and type I error rates for detecting causal SNPs for trait A. For clarity, results are reported as SuperControl minus Standard. Therefore, in the power simulations, positive values indicate better performance of the SuperControl phenotype, while in the type I error simulations, negative values indicate better performance of the SuperControl phenotype due to lower error rates.

The simulations revealed that misclassification and SuperControl bias interact rather than acting separately. For SNPs influencing both trait A and trait B, the SuperControl definition consistently outperformed Standard controls, and this advantage became stronger as misclassification increased (Fig. 2a). For SNPs specific to trait A, Standard controls had more power when misclassification was low. However, as misclassification increased, SuperControl gained power, and at moderate or high levels of misclassification the two approaches yielded similar power for detecting trait A-specific SNPs (Fig. 2b).

**Fig. 2.**
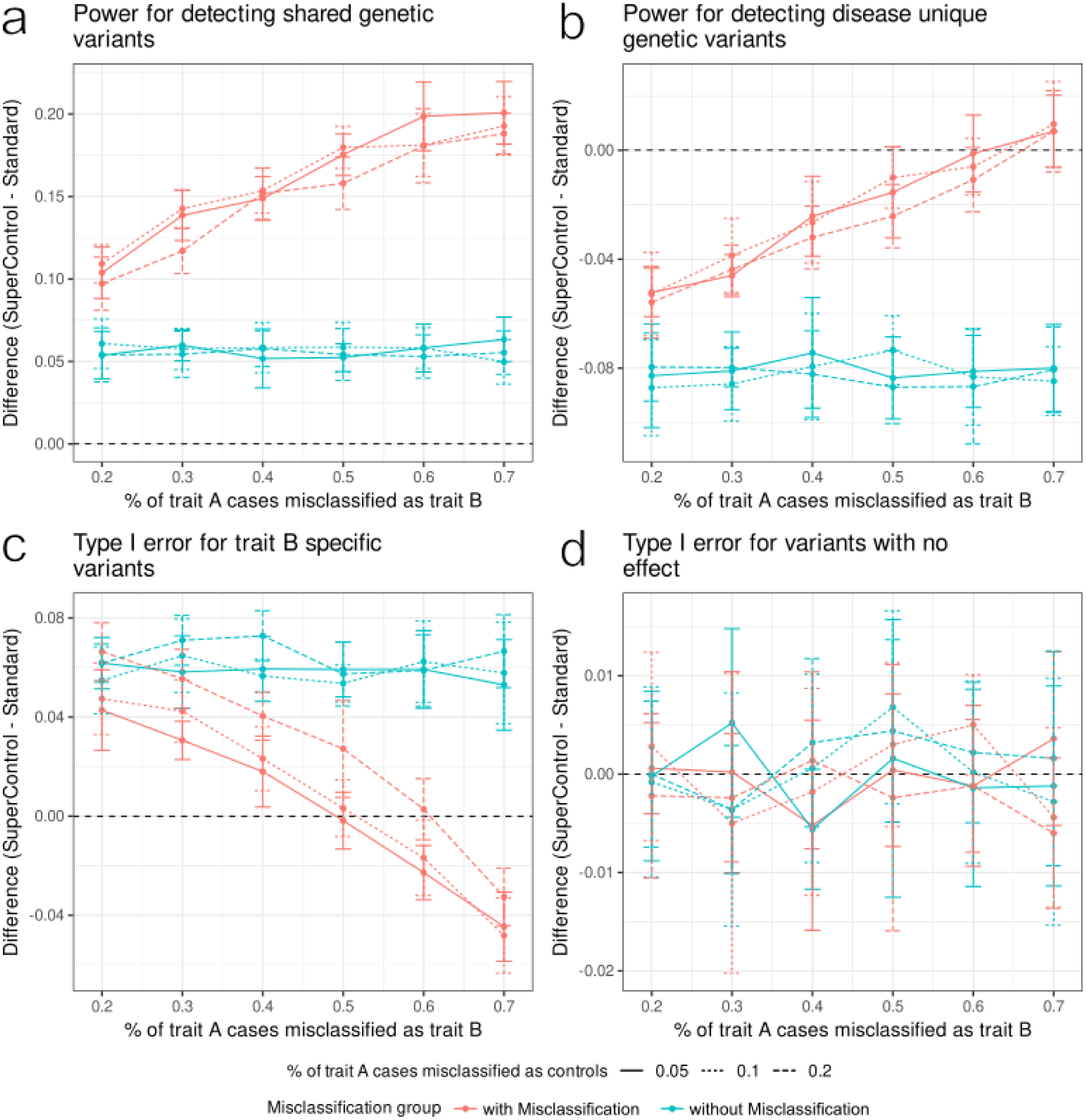
Simulation of SuperControl and Standard phenotypes under varying misclassification at 50% trait prevalence. a–d, Differences (SuperControl – Standard) in power or type I error across increasing levels of misclassification in trait B and among controls other than trait B. a,b, Power for SNPs shared between traits A and B (a) and specific to trait A (b), where positive values indicate better performance of the SuperControl phenotype. c,d, Type I error for SNPs causal only for trait B (c) and for non-causal SNPs (d), where negative values indicate better performance of the SuperControl phenotype due to lower error rates. Curves represent the mean of ten independent simulations, with error bars denoting standard deviation.

We then asked whether SuperControl altered false positive rates for SNPs with no true effect on trait A. For SNPs that affected only trait B, any association with trait A represents a false positive. At low misclassification, SuperControl produced slightly higher error rates than Standard controls. When misclassification reached moderate levels, the two designs performed similarly. With high misclassification, however, the pattern reversed, and SuperControl generated fewer false positives (Fig. 2c). For SNPs with no causal effect on either trait, both definitions performed similarly, with differences in error rates close to zero (Fig. 2d).

Finally, the impact of SuperControl depended strongly on prevalence. When trait A was common (50%), the benefits of SuperControl were most evident (Fig. 2, Supplementary Table 1). At lower prevalence (30% and 10%; Supplementary Figs. 1-2, Supplementary Table 1), differences between SuperControl and Standard, and between data with and without misclassification, became much smaller.

Together, these simulations show that misclassification bias and SuperControl bias interact in predictable ways. The SuperControl approach can recover power and reduce error when misclassification is substantial, particularly for common traits, while providing less added benefit when case definitions are already accurate.

### SuperControl definition improves discovery in FinnGen psychiatric traits despite reduced sample size

We analyzed ten psychiatric traits in FinnGen: obsessive-compulsive disorder (OCD), post-traumatic stress disorder (PTSD), attention-deficit/hyperactivity disorder (ADHD), schizophrenia (SCZ), major depressive disorder (MDD), anxiety disorders (ANX), substance use disorders (SUD), sleep disorders (SLD), bipolar disorder (BD), and a composite trait of any mental health disorder (MHD). Case counts ranged from 2,830 (prevalence 0.6%) for OCD to 59,969 (11.8%) for MDD, with 140,342 (27.6%) for MHD (Supplementary Table 2). These definitions, derived from ICD-9 and ICD-10 codes, are referred to as the Original FinnGen phenotypes.

To reduce misclassification, we constructed a refined control group, the SuperControl phenotype, by excluding all individuals whose psychiatric comorbidities increase the likelihood of being misclassified. After applying these exclusions, 97,181 individuals remained eligible as controls. For each psychiatric trait, the SuperControl phenotype was defined by combining cases from the Original FinnGen phenotype with this refined control set (Supplementary Table 2).

We conducted GWAS for all ten traits using both the Original and SuperControl definitions (Fig. 3; Supplementary Fig. 3). Despite smaller sample sizes, the SuperControl phenotype yielded more genome-wide significant loci, with independence established through conditional analysis. After merging overlapping regions, we identified 94 independent loci for the Original FinnGen phenotype and 201 for the SuperControl phenotype. Of these, 70 were shared between definitions, while 131 were unique to the SuperControl phenotype, representing additional discoveries not captured by the original approach (Table 1).

**Fig. 3.**
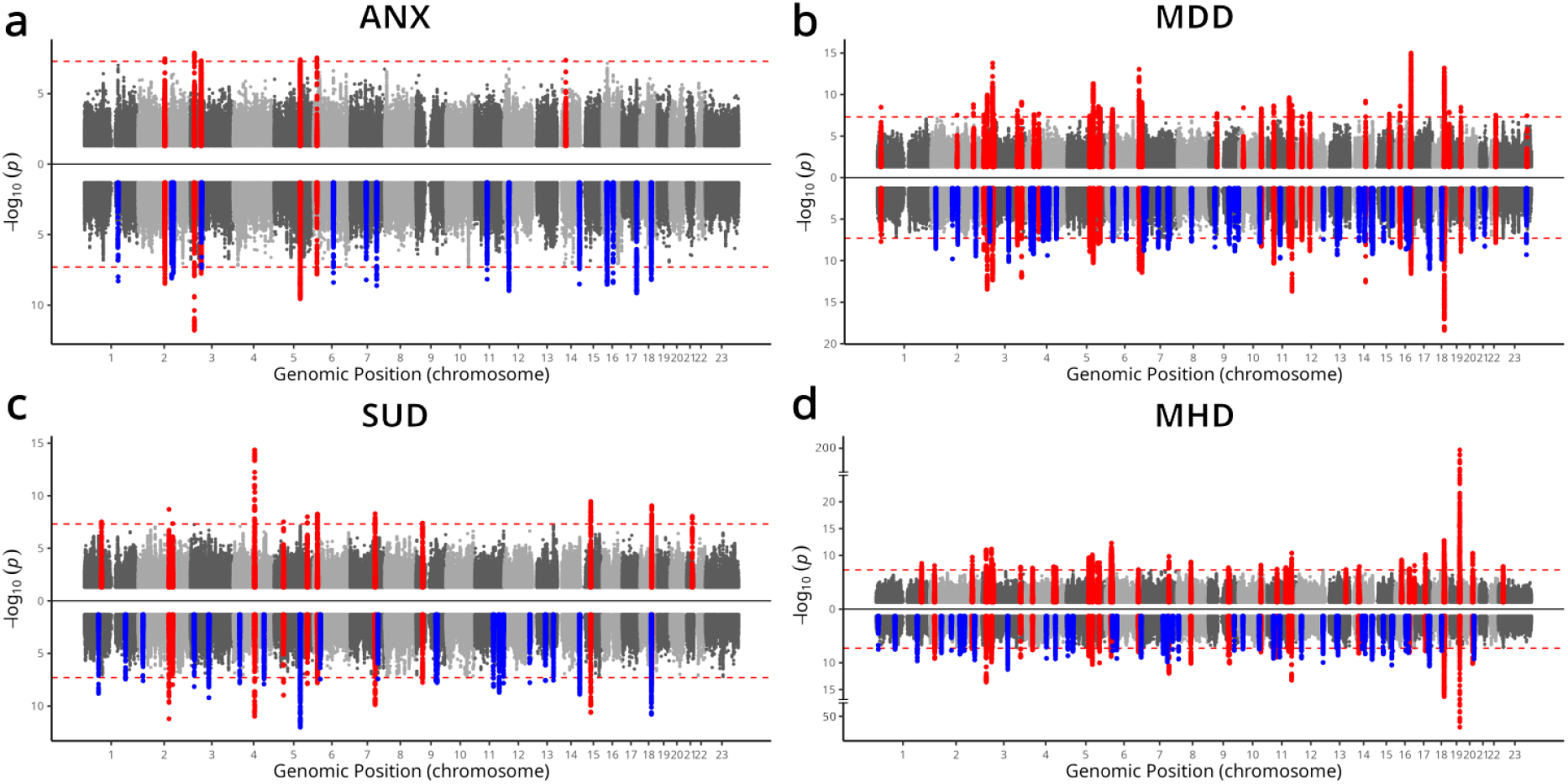
Genome-wide significant loci in Original FinnGen and SuperControl phenotypes across psychiatric traits. a-d, Mirrored Manhattan plots for anxiety (ANX), major depressive disorder (MDD), substance use disorder (SUD), and a combined “any mental health disorder” trait (MHD). In each plot, the upper half represents results from the Original FinnGen phenotype and the lower half from the SuperControl phenotype. Red points denote loci shared between the two phenotypes or unique to the Original FinnGen phenotype, and blue points denote loci unique to the SuperControl phenotype.

The additional discoveries were largely driven by traits with higher prevalence. For less prevalent disorders (OCD, ADHD, SLD, BD; <10,000 cases), the number of additional loci was modest (0-3; Supplementary Fig. 3a-d). In contrast, highly prevalent traits such as ANX, MDD, SUD, and MHD showed a marked increase, with the SuperControl phenotype identifying at least twice as many loci as the Original FinnGen definitions while retaining nearly all original loci (Fig. 3a-d). PTSD and SCZ did not yield genome-wide significant loci under either definition (Supplementary Fig. 3e-f).

To assess the reliability of loci unique to the SuperControl phenotypes, we performed replication analyses using two complementary approaches. **Locus-based replication (LBR)** classified each locus as having *strong prior evidence (SPE)* if a genome-wide significant variant (p < 5 × 10^−8^) was present within a 3 Mb window in the largest external GWAS for the same disorder, *moderate prior evidence (MPE)* if any variant in that region reached p < 5 × 10^−5^, or *no supporting evidence (NSE)* otherwise. **Gene-based replication (GBR)** used the Open Targets variant-to-gene framework to assign the same categories: *SPE* when at least one mapped gene had prior GWAS support, *MPE* when gene-trait links existed through other non-human genetics evidences, and *NSE* when no prior support was found. Loci were considered replicated when supported by either SPE or MPE evidence.

Even the Original FinnGen phenotypes showed modest replication under LBR across the ten disorders (55%), reflecting the general difficulty of replicating psychiatric GWAS across populations. Traits with stronger existing evidence, such as MDD (69%), replicated more consistently than broader or heterogeneous traits like MHD (33%). For SUD, we compared against several large GWAS covering different forms of substance use, yielding a replication rate of 93% and indicating that replication success depends in part on the comprehensiveness of available external data (Supplementary Fig. 4c; Supplementary Table 3). Overall, 57% of loci identified with the SuperControl phenotypes replicated under the LBR approach, increasing the total number of replicated loci from 52 (Original FinnGen) to 114.

Among the 131 loci novel to the SuperControl phenotypes, 69 replicated (53%), indicating that replication rates were not dependent on prior discovery in the Original FinnGen analysis (Fig. 4a; Supplementary Fig. 4a; Supplementary Table 3).

**Fig. 4.**
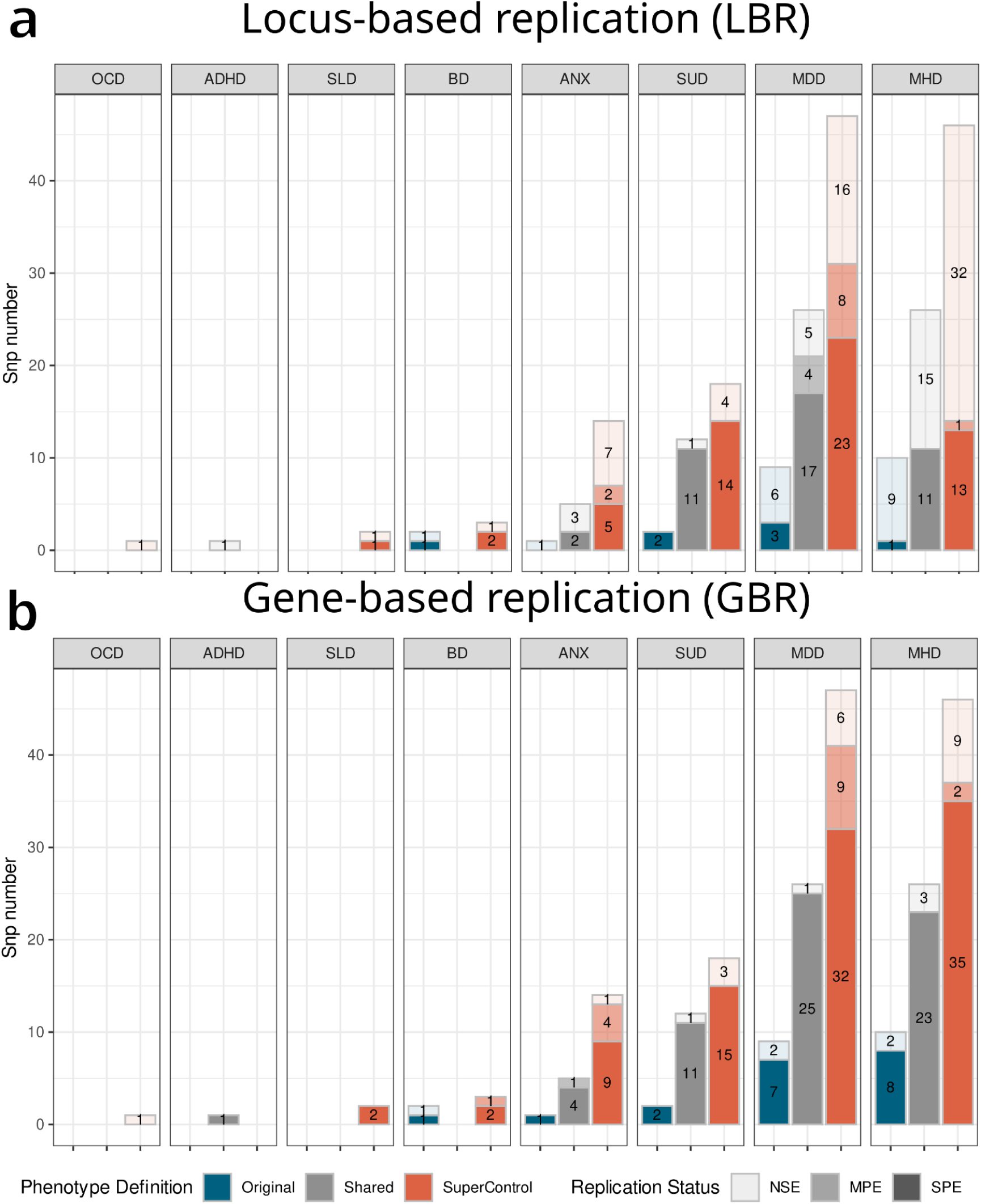
Replication of independent loci in Original FinnGen and SuperControl phenotypes across psychiatric traits. Barplots showing locus-based replication (LBR, a) and gene-based replication (GBR, b) for eight psychiatric traits with genome-wide significant loci: OCD, ADHD, BD, MDD, ANX, SLD, SUD, and the combined “any mental health disorder” trait (MHD). Each trait contains three bars representing loci unique to the Original FinnGen phenotype (left), shared loci between Original FinnGen and SuperControl (middle), and loci unique to the SuperControl phenotype (right). Bars are further segmented by replication strength: strong prior evidence (SPE, darkest shading), moderate prior evidence (MPE, intermediate shading), and no supporting evidence (NSE, faintest shading). SCZ and PTSD are omitted because neither phenotype yielded genome-wide significant loci.

Under the GBR approach, replication rates were higher: 89% for the Original FinnGen phenotypes and 88% for SuperControl. When considering only novel SuperControl loci, 85% replicated at the gene level, demonstrating that most new signals map to biologically relevant genes (Fig. 4b; Supplementary Fig. 4b). Overall, the total number of replicated loci increased from 84 (Original) to 176 (SuperControl) (Fig. 4b; Supplementary Table 4).

Replication rates varied by trait prevalence, with more common disorders showing stronger replication than rarer ones (Supplementary Fig. 4c-d; Supplementary Tables 3,4). This pattern mirrors our simulation results, where SuperControl was most effective for higher-prevalence traits. Importantly, however, direct comparison of definitions showed no significant differences in replication rates (Wilcoxon rank-sum test, LBR: p = 0.28; GBR: p = 0.59; Supplementary Fig. 4e-f), indicating that the additional loci discovered by SuperControl do not come at the cost of reduced reliability.

Together, these findings show that the SuperControl phenotypes substantially increase the discovery of genome-wide loci for psychiatric traits in FinnGen while maintaining replication rates comparable to the Original definitions, despite a smaller sample size.

### SuperControl preserves genetic distinctiveness while enhancing biological specificity

After confirming that SNP-level replication is preserved for the SuperControl phenotype, one central concern remained. By excluding individuals with comorbid disorders the SuperControl approach might artificially increase genetic similarity between related psychiatric traits. To test this, we examined whether refining control definitions altered the overall pattern of genetic relationships across traits. In parallel, we assessed whether the SuperControl design maintained trait-specific biological signatures by comparing tissue-specific heritability enrichment.

We estimated pairwise genetic correlations across all ten psychiatric traits using both the Original FinnGen and SuperControl phenotypes (Fig. 5 a,b; Supplementary Tables 5). The overall structure of genetic relationships was highly preserved between the two definitions, with pairwise correlations showing strong concordance (Pearson r = 0.96, p < 2.2 × 10⁻¹⁶). However, mean pairwise genetic correlations across all ten psychiatric traits were moderately higher under the SuperControl definition (mean difference = 0.19, p = 6.5 × 10⁻¹³), indicating a small but systematic increase in shared genetic signal. Trait pairs with *low* baseline genetic correlations show proportionally high increase in genetic correlations when considering SuperControl phenotypes vs Original FinnGen, whereas highly correlated pairs showed minimal divergence (Supplementary Fig. 5 a).

**Fig. 5.**
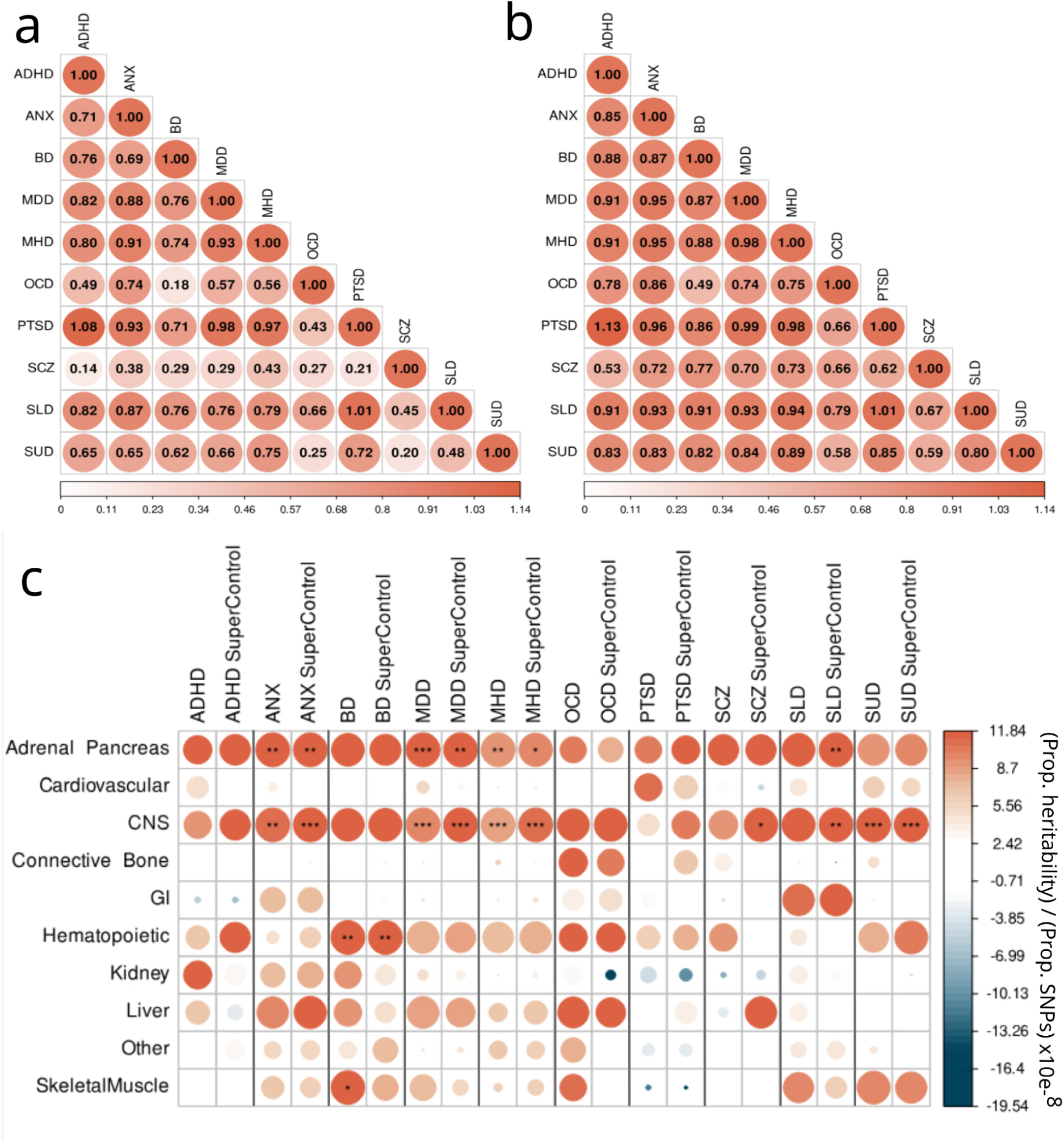
Genetic architecture improvements through refined phenotyping. All panels show data for the 10 psychiatric traits (OCD, PTSD, ADHD, SCZ, MDD, ANX, SUD, SLD, BD, and the combined “any mental health disorder” trait, MHD). (a,b), Heatmaps of pairwise genetic correlations among the traits. (a) shows correlations for the Original FinnGen phenotypes, and (b) for the SuperControl phenotypes. (c), Heatmap of partitioned heritability using the cell-type baseline dataset. Columns represent the 10 traits, and rows represent cell types. Stars indicate significant enrichment.

To examine how these changes affected genetically-informed tissue-enrichment, we compared partitioned heritability coefficients across cell-type-specific annotations between the two Phenotype definitions (Supplementary Table 6). Across ten psychiatric traits, enrichment patterns were largely similar between the Original and SuperControl definitions, with most tissues showing no significant differences after multiple-testing correction. The strongest change was observed for the central nervous system (CNS), where enrichment increased significantly under the SuperControl definition (p = 1.9 × 10⁻³; FDR-adjusted p = 0.02; Supplementary Fig. 5b), consistent with a more brain-specific genetic signal. Nominal decreases were observed for peripheral tissues, including kidney (p = 0.02), cardiovascular (p = 0.04), and skeletal muscle (p = 0.04), although these did not remain significant after multiple-testing correction. Together, these results suggest that the SuperControl definition strengthens CNS-specific enrichment while reducing residual signal in peripheral tissues (Supplementary Table 7).

Trait-level analyses supported this pattern. Under the SuperControl definition, CNS enrichment became significant for several disorders that lacked such evidence in the Original FinnGen phenotypes, including schizophrenia (p = 1.7 × 10⁻²), and sleep disorders (p = 3.8 × 10⁻³). Conversely, nominally significant skeletal muscle enrichment previously detected for bipolar disorder (Original p = 2.0 × 10⁻²) was no longer significant (p = 4.3 × 10⁻¹), aligning with the overall reduction of peripheral enrichment. Notably, for sleep disorders, adrenal-pancreas enrichment reached significance (p = 7.6 × 10⁻³) despite no consistent signal in other traits (Fig. 5 c). This finding aligns with the established role of endocrine pathways, such as cortisol and other hormonal rhythms, in regulating sleep-wake cycles^16^, suggesting that refined control definitions improve detection of biologically coherent, trait-specific signals.

Together, these results demonstrate that the SuperControl approach modestly increases correlations between psychiatric traits but preserves their overall genetic distinctiveness. At the same time, it enhances biological specificity, strengthening enrichment for brain-related pathways and uncovering disorder-relevant signals such as adrenal-pancreas involvement in sleep regulation.

### Integrating excluded individuals via PRISMA restores GWAS power with modest trade-offs in replication and specificity

The SuperControl design improved discovery by a cleaner phenotype definition, but its main limitation was the exclusion of up to 50%-80% of the controls, substantially reducing statistical power. To address this trade-off, we asked whether the excluded individuals could be reintegrated through predictive modelling of their probability of being disease cases, without re-introducing misclassification bias.

PRISMA (Predicted Excluded Individuals and SuperControls via MTAG-based Analysis) enables the recovery of excluded individuals while preserving the advantages of refined control definitions. In the first step, we trained gradient-boosted tree models to predict the SuperControl phenotypes. To do so, we included 4,567 disease traits, 531 medication variables, and additional demographic and lifestyle covariates. Model performance was high across traits (AUC = 0.87-0.98; Supplementary Figs. 6-15). The trained models were then applied to individuals excluded from the SuperControl phenotype definitions to estimate their case probabilities. For all traits except OCD, the predicted probabilities were bimodal, with clear peaks near 0 and 1, indicating strong separation between cases and controls. Next, we performed GWAS on binarized predicted probabilities using a threshold of 0.5. To reiterate, these GWAS were conducted only among individuals not used to define the SuperControl phenotypes. For most traits, genetic correlations between the SuperControl and excluded-cohort GWAS ranged from 0.6 to 0.7 (Supplementary Fig. 16, Supplementary Table 8). Finally, the GWAS results from the SuperControl phenotypes and from the binarized predicted probabilities were combined using MTAG (Supplementary Fig. 17).

Leveraging the full FinnGen cohort via PRISMA led to a clear increase in genome-wide discovery compared with both the SuperControl and Original phenotypes (Fig. 6a; Supplementary Fig. 4a-b). After merging overlapping regions, we identified 307 independent loci for PRISMA, compared with 201 for the SuperControl and 94 for the Original FinnGen phenotypes. Of these, 168 loci were shared between PRISMA and SuperControl, while 139 loci were unique to PRISMA, and only 33 remained unique to SuperControl (Table 1). Thus, PRISMA increased discovery by 1.5-fold relative to SuperControl and threefold relative to the Original FinnGen phenotypes.

**Fig. 6.**
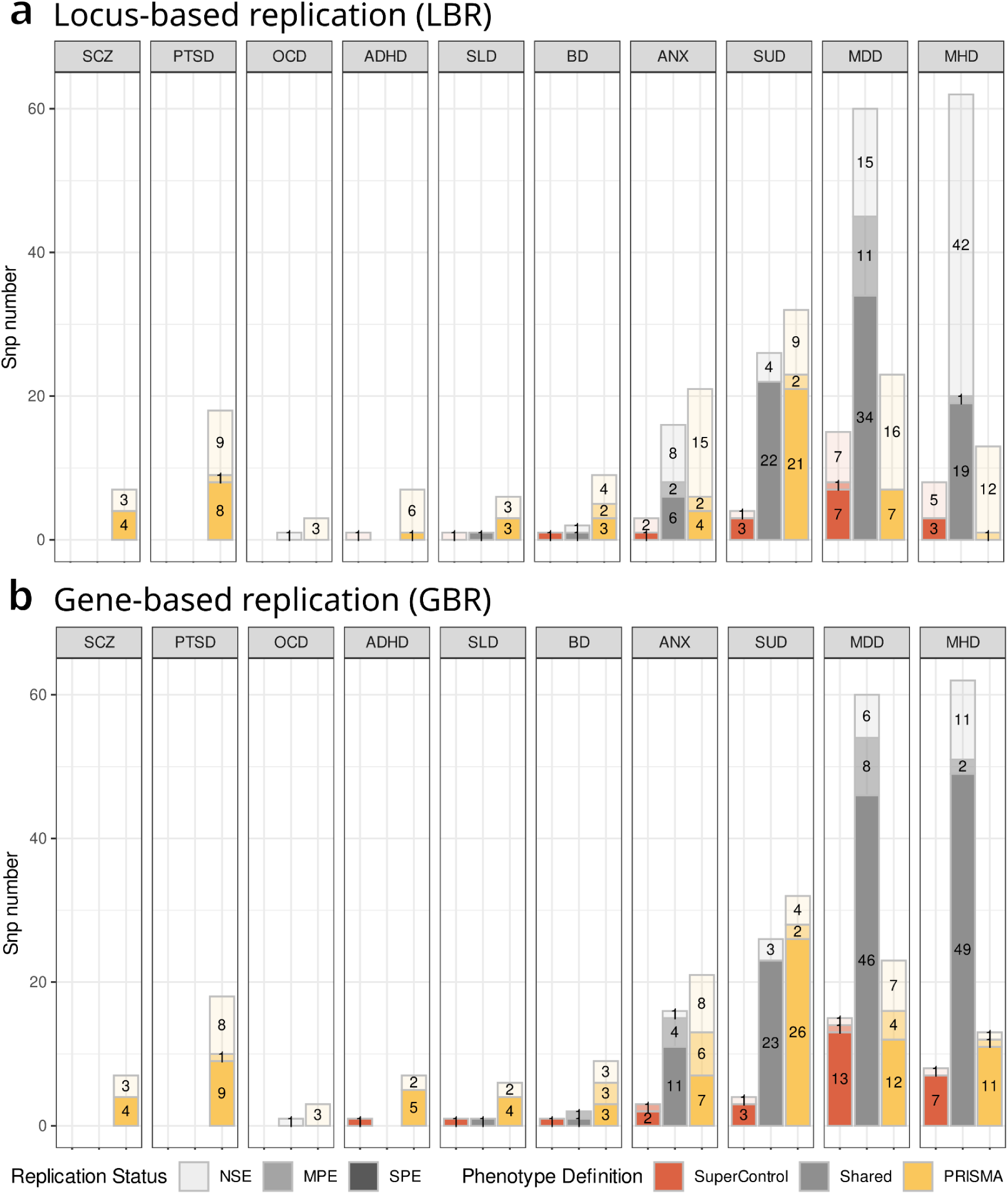
Replication of independent loci in SuperControl and PRISMA phenotypes across psychiatric traits. a-b Barplots showing locus-based replication (LBR, a) and gene-based replication (GBR, b) for ten psychiatric traits: SCZ, MDD, BD, ANX, ADHD, PTSD, OCD, SLD, SUD and MHD. Each trait contains three bars representing loci unique to the SuperControl phenotype (left), shared loci between SuperControl and PRISMA (middle), and loci unique to the PRISMA phenotype (right). Bars are further segmented by replication strength: strong prior evidence (SPE, darkest shading), moderate prior evidence (MPE, intermediate shading), and no supporting evidence (NSE, faintest shading).

While gains were modest for the two largest traits (MDD and MHD; > 50,000 cases), likely reflecting saturation of detectable signal at current GWAS resolution, PRISMA revealed new associations for previously underpowered disorders. Notably, PTSD and schizophrenia, which had no significant loci under either previous definition, reached genome-wide significance in PRISMA (Supplementary Fig. 4a-b, 17 i,j). For PTSD and schizophrenia, we identified 18 and 7 genome-wide SNPs in the PRISMA phenotypes, respectively. Of these, 8 PTSD SNPs and 4 schizophrenia SNPs reached genome-wide significance in the largest published GWAS for each disorder (*Nievergelt et al.*, 2024^17^; *Trubetskoy et al.*, 2022^18^). Gene annotation using Open Targets revealed that several PRISMA loci overlapped with genes previously found in the PTSD GWAS of *Nievergelt et al.* PTSD meta-analysis, including *FOXP2, DCC, ESR1, LRRC37A2, ARHGAP27*, and *C17orf58*. Notably, *DCC* was identified as a genome-wide significant gene in both the *Nievergelt et al.* PTSD study and the *Trubetskoy et al.* schizophrenia GWAS, and was also detected in the PRISMA phenotypes for both disorders, marking it as a strong cross-disorder signal.

Despite a higher number of discoveries, replication performance remained broadly similar across phenotypes. Locus-based replication (LBR) rates were 55% for Original FinnGen, 57% for SuperControl, and 51% for PRISMA, while gene-based replication (GBR) rates were 89%, 88%, and 79%, respectively. However, the absolute number of replicated loci increased with PRISMA, from 52 (Original) and 114 (SuperControl) to 156 (LBR) and from 84 (Original) and 176 (SuperControl) to 244 (GBR) (Fig. 6a-b; Supplementary Tables 3,4).

Gains were most evident for intermediate-sized traits, such as sleep disorders, bipolar disorder, PTSD, and schizophrenia, where improved prediction offset the reduced power of the SuperControl design. In contrast, traits with limited training signal, such as OCD, yielded few discoveries and no replication (Supplementary Figs. 6-15).

Genetic correlations remained broadly consistent with the Original definitions but showed slightly higher cross-trait sharing, indicating modest inflation of pleiotropy under the imputed phenotypes (Supplementary Fig. 18a-b). Partitioned heritability analyses showed that CNS enrichment was largely maintained but somewhat reduced relative to SuperControl, consistent with a partial loss of biological specificity (Supplementary Fig. 18c-d; Supplementary Tables 6,9).

Overall, PRISMA recovered signals consistent with known loci while expanding discovery across several psychiatric traits. Its advantages were most pronounced for traits with intermediate prevalence, where it restored power lost under stricter control definitions, whereas gains were smaller for large, well-powered traits and unreliable for those with few cases. These results indicate that PRISMA complements the SuperControl framework, enhancing discovery and replication while maintaining broad biological consistency.

### SuperControl and PRISMA-derived polygenic scores are more predictive of mental health disorders in an external study

After demonstrating stronger discovery within FinnGen, we next tested whether these refined genetic signals generalize to external cohorts. Specifically, we evaluated how well polygenic scores (PGS) derived from the Original, SuperControl, and PRISMA GWAS predict psychiatric traits in UK Biobank (UKBB).

PGS from all three Phenotypes were evaluated for ten ICD-10-defined psychiatric traits in UKBB (N = 408,792). However, only four traits had more than 2,000 cases: anxiety (ANX; 33,494 cases), depression (MDD; 48,160), substance use disorder (SUD; 25,929), and any mental disorder (MHD; 96,478). Across these, all PGSs showed significant associations with the traits (e.g., ANX: p = 1.3 × 10⁻152; MHD: p = 0). For the remaining low-prevalence traits (<2,000 cases), estimates were less stable (Supplementary Fig. 19; Supplementary Tables 10,11).

The SuperControl PGS consistently outperformed the Original FinnGen scores, achieving significantly higher odds ratios for ANX (p = 3.6 × 10⁻²), MDD (p = 5.4 × 10⁻³), MHD (p = 9.8 × 10⁻⁸), and SUD (p = 6.4 × 10⁻³). Similarly, PRISMA PGS improved prediction for MDD (p = 3.3 × 10⁻³), MHD (p = 7.2 × 10⁻¹⁰), and SUD (p = 2.9 × 10⁻⁸) compared to the Original FinnGen phenotype (Fig. 7a; Supplementary Table 10). Together, these findings confirm that refined phenotyping enhances out-of-sample prediction performances of polygenic scores.

**Fig. 7.**
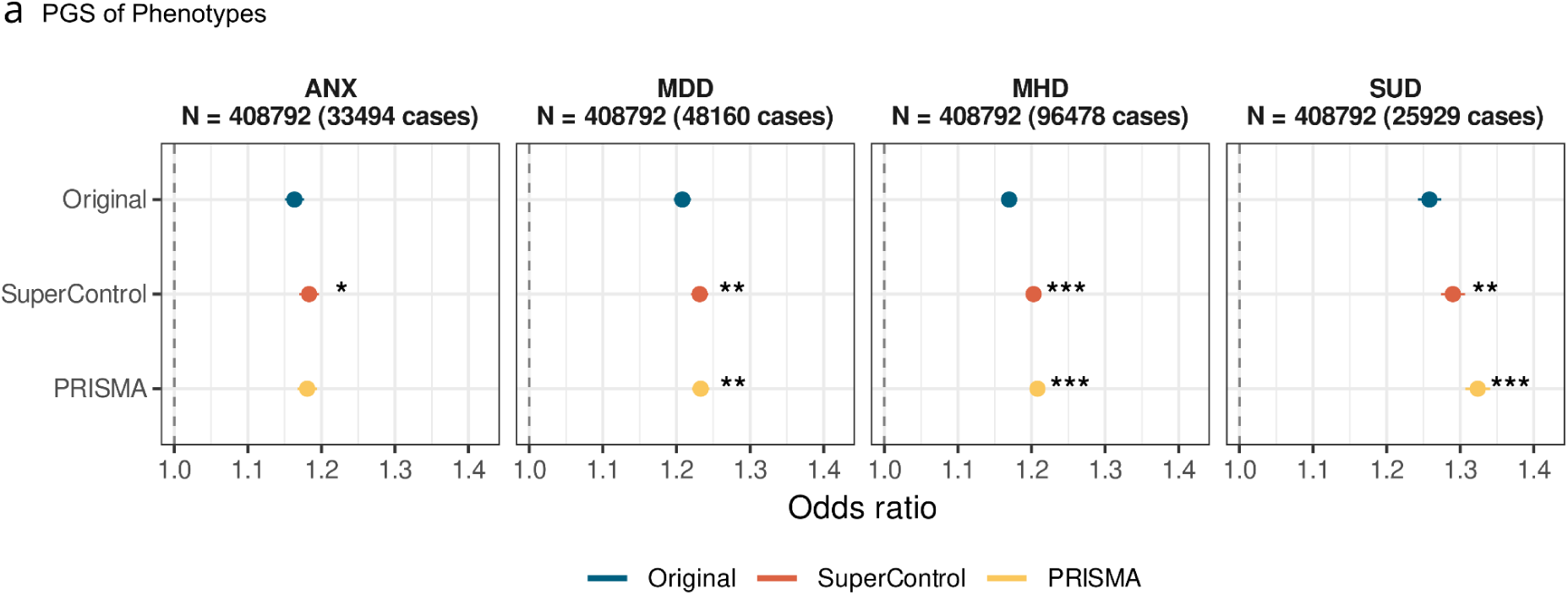
Polygenic prediction across biobanks. a, Odds ratios (with standard errors) from polygenic score (PGS) prediction for four psychiatric traits, ANX, MDD, MHD, and SUD, based on GWAS from the Original FinnGen, SuperControl, and PRISMA phenotypes. Asterisks indicate traits where the SuperControl or PRISMA PGS showed significantly stronger prediction than the Original FinnGen PGS.

To assess whether non-replicated loci still contributed informative signal, we constructed PGS restricted to lead SNPs that did not replicate in external GWAS using LBR. For each such lead SNP, we included all variants within a ±1.5 Mb window to capture local linkage structure. This stringent filtering substantially reduced the number of variants and, as expected, led to markedly attenuated prediction accuracy (ΔR² = 0.00003-0.001). Despite this reduction, the resulting PGS remained predictive of well powered UKBB traits. This demonstrates that even loci, that did not replicate according to our criteria, contain trait-relevant genetic signal (Supplementary Fig. 19c; Supplementary Table 11).

## Discussion

Large-scale case-control GWAS in psychiatric genetics have advanced rapidly in recent years, yet one foundational issue remains unresolved: how best to define controls.

Historically, the use of “super-normal” controls (i.e. excluding individuals with psychiatric comorbidities) was introduced to create a deeper phenotype and reduce noise in early, small-scale studies where statistical power was limited^11,19^. However, this strategy quickly drew criticism due to its potential to introduce selection bias and inflate genetic signal by reducing trait variance in the control group. Thoughtful critiques by Schwartz & Susser highlighted how “super normal” controls could distort effect sizes and reduce generalizability in both epidemiological and genetic studies^13,14^. As larger, uncurated datasets, particularly those derived from EHR, became available, many studies shifted toward using “unscreened” population-based controls instead (*shallow phenotype*).

Yet this shift introduced a new problem: misclassification bias^6^. Psychiatric phenotypes, especially those with high comorbidity, underdiagnosis, and social stigma, are prone to diagnostic inaccuracies in EHR-based data. Recent work by Hodge et al. and Kendler et al. has demonstrated that both unscreened and “super normal” controls introduce different forms of bias, and that excluding ambiguous or potentially misclassified individuals may, improve power and reduce distortion even at the cost of smaller sample sizes^6,20^. However, prior studies have examined missingness bias and control-definition bias separately, leaving the interaction between these biases largely unexplored.

To address this gap, we developed the SuperControl phenotype, a data-driven, generalized adaptation of the classic “super normal” control concept. This approach excludes individuals most likely to be misclassified cases or affected by structured missingness in the EHR data, aiming to reduce noise while maintaining sample validity.

Simulations showed that the SuperControl approach restores power lost when traits are missing or misclassified. It improves detection of shared genetic signals between related disorders while keeping similar power for trait-specific effects. Under high levels of missingness, SuperControls also reduced false positives for trait-specific signals compared to standard definitions, without inflating signals unrelated to the analyzed or control-defining traits. These benefits were strongest for traits with higher prevalence, both in the target disorder and in those contributing to the SuperControl group. Overall, this suggests a simple mechanism: by excluding individuals most likely to be misclassified, the SuperControl design removes noise from undiagnosed or uncertain cases, allowing true genetic associations to appear more clearly.

We next assessed this principle in the FinnGen study, which already incorporates some medical curation in phenotype definitions. Even in this relatively well-curated setting, applying the SuperControl phenotype improved discovery for several major psychiatric traits. To characterize these effects more fully, we evaluated genetic signal, replication, cross-biobank transferability, and tissue-specific heritability across ten major psychiatric traits.

Despite a reduction in cohort size, the SuperControl phenotype led to a marked increase in genome-wide discovery, with up to double the number of significant loci for several traits. Notably, replication rates remained comparable to those observed using the original FinnGen phenotype, echoing previous findings that filtering out potential misclassification can improve signal without degrading validity^6^. These results suggest that using the SuperControl Phenotype increases signal discovery and power of psychiatric GWAS.

While we acknowledge the long-standing critique that super normal controls may inflate effect sizes or obscure trait boundaries, we found evidence of maintained genetic distinction between traits using the SuperControl phenotype. For example, partitioned heritability analyses revealed consistent trait-specific enrichment patterns, with subtle shifts suggesting both improved signal and some blending of comorbid dimensions. Genetic correlations between traits were slightly increased, particularly for schizophrenia, with the overall pattern still faintly visible.

Biologically, we observed meaningful differences in tissue-specific heritability signals across phenotype definitions. The SuperControl phenotype showed stronger enrichment in central nervous system (CNS) tissues and reduced signal in peripheral tissues, suggesting greater biological specificity. Enrichment patterns in adrenal-pancreas tissues also revealed notable trait-specific distinctions: for anxiety and depression, pancreatic enrichment was consistently detected in both the original and SuperControl phenotypes, which may reflect plausible endocrine contributions to mood-related symptoms^21,22^. In contrast, sleep disorder showed a clear enrichment in the SuperControl phenotype, absent in the original FinnGen definition, for adrenal-pancreas tissues and CNS, which is particularly notable given the well-documented links between endocrine and metabolic pathways and sleep regulation^16^. This suggests that the SuperControl phenotype may enhance biological specificity by filtering out misclassified cases, allowing clearer detection of trait-relevant signals.

Several studies have recently trained predictive models to address missingness in EHR-based studies, including imputing diagnoses^7,23^, broadening narrow phenotypes^2^, incorporating additional data sources such as clinical records^9,24,25^ or family history^26,27^, applying liability-scale modeling^28^, or predicting disease onset^29^. Despite these advances, important limitations remain. First, some approaches rely on questionnaire data, clinical assessments, or family history, requiring information that is not routinely available in most EHR systems. Second, several methods report novel loci that may reflect prediction noise rather than genuine biological signal. While some studies attempt to mitigate this by filtering loci against GWAS of the original phenotype^28^, this strategy implicitly assumes that the underlying EHR-derived phenotype is accurate, an assumption that is problematic given well-documented misclassification and bias in real-world health data. Third, many models assume missingness at random, whereas in EHR-based cohorts missing data is often systematically structured by selection and participation biases, including access to care, diagnostic practices, and healthcare system dynamics^30^.

Building on this prior work, and leveraging the strengths of the SuperControl framework, we developed PRISMA, a machine-learning approach that imputes disease liability probabilities among individuals excluded by the SuperControl definition. GWAS of these imputed liabilities were then combined with the SuperControl GWAS using MTAG. While PRISMA introduced additional noise, it occasionally outperformed the SuperControl GWAS, particularly for traits with larger case counts or those prone to underdiagnosis, such as substance use disorders, anxiety, and depression. In contrast, PRISMA was less effective for traits with fewer cases or greater phenotypic heterogeneity. Although phenotype imputation requires caution due to the risk of amplifying existing biases, our results suggest that the additional signal captured by PRISMA is biologically meaningful. Notably, PRISMA identified genome-wide significant loci for PTSD and schizophrenia that were not detected using the original or SuperControl phenotypes. Many of the PTSD-associated genes overlapped with findings from previous large-scale GWAS, implicating pathways involved in neuronal development, hormone signaling, and stress response^17^. Key examples include *FOXP2*^31,32^, *DCC*^33^ (neural circuit formation) and *LRRC37A2*^34,35^ (neuronal and stress-related processes). Moreover, the PRISMA phenotype demonstrated clearer advantages in cross-cohort prediction, with significant improvement of disease prediction using PGS compared to Original phenotypes and SuperControl.

More broadly, our study also touches on pressing issues in the design of EHR-based genetic studies. Recent evaluations have shown that cohort-level variation (e.g., biobank, country, ancestry) can exceed method-level variation in PGS performance, reinforcing the need for international harmonization^36^. Furthermore, structural biases in healthcare access, recruitment, and diagnosis, especially for socially stigmatized conditions like psychiatric disorders, are increasingly recognized as major sources of distortion in both GWAS and predictive modeling^3–5^. Machine learning approaches, if not carefully evaluated, risk learning and amplifying those biases, particularly when missingness is non-random and diagnoses reflect social inequities rather than biological variation.

Despite these challenges, our findings highlight a practical and scalable strategy to address one of the field’s most persistent issues: control misclassification in GWAS using real-world data. By systematically comparing unscreened, SuperControl, and predicted phenotypes, we demonstrate that thoughtful phenotype refinement can enhance power, maintain replicability, and yield biologically meaningful insights, even within the inherently noisy context of EHRs. This approach is particularly well-suited for traits with sufficient case numbers, where the robustness of the signal can offset the modest noise introduced by stricter control definitions or predictive methods. While this approach increases pleiotropy, our simulations demonstrate that misclassification bias and the unspecific signal introduced by the SuperControl phenotype interact, and that under certain conditions the benefits of reducing misclassification outweigh the modest increase in pleiotropy. Based on conceptual simulations showing reduced performance for low-prevalence traits, we recommend applying the SuperControl framework primarily to more prevalent traits, balancing improved power against control exclusion rates.

## Methods

### Cohort Information: FinnGen and UK Biobank

We analyzed data from two population-scale cohorts, the FinnGen study (Data Freeze 12) and the UK Biobank (UKBB), both of which link genome-wide genotyping data to longitudinal health records. FinnGen includes 520,149 anonymized participants, representing approximately 10% of the Finnish adult population. The cohort is enriched for disease traits and integrates genetic data with nationwide health registries, including hospitalizations, outpatient visits, cancer diagnoses, medication purchases, and demographic variables such as smoking status, occupational history, and number of children. The median age of participants at the time of sample collection was 53 years, with 43% male and 57% female^37^. UKBB includes approximately 500,000 anonymized individuals with linked genetic data and comprehensive EHR, including primary care data, diagnoses, procedures, and prescriptions ^38,39^. Genotyping in both cohorts was performed using arrays with 700,000-800,000 markers per individual. Disease traits were harmonized using International Classification of Diseases, 10th Revision (ICD-10) codes to enable cross-cohort comparisons. Detailed information on genotyping procedures and cohort composition for both UK Biobank and the FinnGen study is provided in the Supplementary Methods.

### Simulation Framework for Evaluating Supernormal Design and Misclassification in GWAS

To investigate the impact of using a supernormal design and the presence of misclassified controls in genome-wide association studies (GWAS), we conducted simulations incorporating both factors. We modeled two genetically overlapping traits, A and B, in a cohort of N=10,000 individuals, where traits A represents the traits of interest for GWAS, and trait B is to be taken into account for control inclusion in a supernormal design.

We simulated four groups of single nucleotide polymorphisms (SNPs):

1. k_1_ = 500 SNPs with shared causal effects on both trait A and B;
2. k_2_ = 500 SNPs with causal effects only on trait A;
3. k_3_ = 500 SNPs with causal effects only on trait B;
4. k_4_ = 500 SNPs with no causal effects on either trait.

Genotypes for all SNPs were simulated independently with a minor allele frequency of 0.3. For each individual *i*, liability for a trait was modeled as:

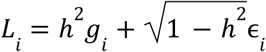

Where ℎ^2^ = 0.6 is the trait heritability, *g_i_* is the individual’s genetic risk and ɛ*_i_* ∼ *N*(0, 1) represents environmental noise. For genetic risk *g_i_*, we first generated individual *i*’s raw genetic risk 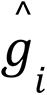 as the aggregated polygenic risk of all *k* = 1,000 causal SNPs for the trait (For trait A: *k_A_* = *k*_1_ + *k*_2_, for trait B: *k_B_* = *k*_1_ + *k*_3_).

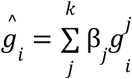

Where *β_j_* ∼ *N*(0, 1) is the effect size of causal variant *j*, and *g*^*j*^_*i*_ ∈ { 0, 1, 2 } is the simulated genotype coded as the number of effective alleles for individual *i* at causal variant *j*. Once raw genetic risk was generated for all individuals, we scaled 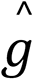 across the whole cohort to get *g_i_* so that *g_i_* ∼ *N*(0, 1).

After creating the liabilities, we then used a liability threshold model to assign the trait diagnosis *T_i_* for an individual _i_ as below

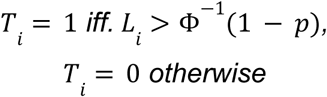

where Φ(*) is the cumulative density function of a standard normal distribution, and *p* is the prevalence of the trait. We set the prevalence for both trait A and B to be the same and varied the value as below

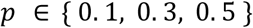

After generating binary diagnoses for both traits, we introduced misdiagnosis into the labels for trait A. Specifically, we defined two proportions:

- *P_B0_*: the proportion of individuals with A=1, B=0 who were misclassified as A=0 (i.e., undiagnosed);
- *P_B1_*: the proportion of individuals with A=1, B=1 who were similarly misclassified as A=0.

We assumed *P_B1_* ≥ *P_B0_*, reflecting expectations in real-world scenarios for highly comorbid traits such as psychiatric disorders. These proportions were varied as follows:

These proportions were varied as follows:

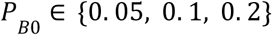

and

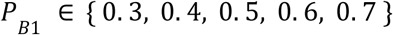

Under this setup, we performed four GWAS analyses for trait A using logistic regression:

1. Standard GWAS without misdiagnosis, where cases are defined as individuals with A = 1 and controls are defined as individuals with A = 0, using original labels;
2. Supernormal GWAS without misdiagnosis, where cases are defined as individuals with A = 1 and controls are defined as individuals with A = 0 and B = 0, using original labels;
3. Standard GWAS with misdiagnosis, where cases are defined as individuals with A = 1 and controls are defined as individuals with A = 0, using labels with introduced misdiagnosis;
4. Supernormal GWAS with misdiagnosis, where cases are defined as individuals with A = 1 and controls are defined as individuals with A = 0 and B = 0, using labels with introduced misdiagnosis.

We evaluated each GWAS using the following four performance metrics: a. power to detect SNP with shared causal effect on both trait A and B (k_1_); b. power to detect SNPs with causal effect specific to trait A (k_2_); c. type I error in picking up SNPs with causal effect specific to trait B (k_3_); and d. type I error in picking up non-causal SNPs (k_4_). All evaluations were based on a significance threshold of p<0.05. All simulations were repeated for 10 independent iterations, and we reported the mean and standard deviation of each metric of interest.

### Super Control Phenotype and traits

This study focused on ten major psychiatric disorders: sleep disorders (SLD), attention-deficit/hyperactivity disorder (ADHD), bipolar disorder (BD), schizophrenia (SCZ), obsessive-compulsive disorder (OCD), post-traumatic stress disorder (PTSD), anxiety (ANX), depression (MDD), substance use disorder (SUD), and a composite "any mental health disorder" trait (MHD) aggregating all mental diagnoses. These disorders were selected based on their substantial public health impact, chronic course, and high rates of underdiagnosis, as well as their varying prevalence and degrees of prior characterization in the literature. For the reference GWAS we use the original FinnGen Phenotype. These phenotypes are constructed using International Classification of Diseases (ICD) codes recorded in patients’ EHR, and curated by the FinnGen clinical team. Case definitions typically rely on one or more relevant diagnostic codes, while the remaining population is used as controls. In some cases, control definitions are refined by excluding individuals with highly comorbid diagnoses. Links to Phenotype definitions can be found in Supplementary Table 2.

For the Super Control Phenotype we excluded all individuals with any diagnosed psychiatric trait from the controls, as psychiatric disorders are known to exhibit high comorbidity with one another^40^. We also excluded all individuals with neurodegenerative diseases, given the documented comorbidity and suspected shared biological mechanisms^41^. To further ensure the removal of individuals with likely underlying psychiatric conditions, we additionally excluded participants with records indicating the use of one of the 53 psychiatric or neurologically active medications (All excluded traits can be found in Supplementary Table 3).

To minimize diagnostic overlap and potential misclassification, we defined a stringent control group (“Super Controls”) for each disorder. Cases were defined as individuals with at least one diagnosis code corresponding to the disorder of interest. Controls were excluded if they had any diagnosis of a mental or neurodegenerative disorder (n = 489 traits) or any prescription for psychiatric medications (n = 53 drug categories, identified using Anatomical Therapeutic Chemical [ATC] classification codes)(Supplementary Table 3).

Following these exclusion criteria, the final control set comprised 97,181 (90,667) individuals. Case counts vary by disorder and are reported in Supplementary Table 2.

### PRedicted excluded individuals and Super Control cohort integrated via MTAG Analysis (PRISMA)

Prior to model training, traits and covariate data were preprocessed to ensure consistency and relevance. All non-core traits were excluded, and any remaining events prior to 1995 were removed. For each retained trait, the year of first occurrence and the total number of events were included as features. Medication variables were derived from Anatomical Therapeutic Chemical (ATC) codes and supplemented with VNR package-level data, enabling the estimation of total prescriptions, package sizes, and approximate intake counts. Socioeconomic covariates included the highest attained educational degree and corresponding field of study, based on Statistics Finland’s classification of education levels. Occupational categories were assigned using the national classification of occupations.

To infer psychiatric risk among individuals excluded from the Super Control cohort, we trained a supervised machine learning model using gradient boosted decision trees implemented via XGBoost in Python. The input features included 4,567 binary disease traits, 531 medication variables, and demographic and lifestyle covariates (education level, occupational field, region of birth, smoking status, and number of children). All traits and medication variables used in the construction of the Super Control cohort were excluded from the feature set to ensure that the model did not rely on variables that were, by design, absent in the control group and inherently linked to case status.

Model training was performed exclusively within the Super Control cohort. The dataset was randomly split into 80% training and 20% validation sets, stratified by case-control status. A nested split within the training set (20%) was used for hyperparameter optimization via Bayesian optimization using BayesSearchCV from the scikit-optimize package. The final model was retrained on the full training set using the best-performing hyperparameters and evaluated on the held-out validation set (Supplementary Fig. 6-15 a-d).

After model training, we applied the fitted classifier to individuals in the excluded cohort. Model outputs represent predicted probabilities of psychiatric disease liability. Based on the observed bimodal distribution of prediction scores (Supplementary Fig. 6-15e), we applied a binary classification threshold of 0.5 to define high-risk individuals. Individuals above this threshold were labeled as "predicted cases" and used to construct a synthetic case group within the excluded cohort.

To increase discovery power while accounting for genetic correlation between observed and predicted phenotypes, we applied Multi-Trait Analysis of GWAS (MTAG; accessed August 2024)⁴ to jointly analyze summary statistics from the Super Control phenotype and the predicted phenotype in the excluded cohort. This approach yielded a combined phenotype, termed **PRISMA** (PRedicted excluded Individuals and Super Controls via MTAG-based Analysis).

We used LD score regression (LDSC v1.0.1)^42^ to estimate genetic correlations between the Super Control and excluded cohort phenotypes. Most traits showed moderate to high genetic correlation (rg = 0.6-0.7), with all estimates exceeding 0.3, a level previously considered sufficient (Munio et al)⁶ for joint analysis in large-sample GWAS^43^ (see Supplementary Fig. 15).

### Bias Considerations

Because the SuperControl Phenotype was constructed by excluding individuals with known psychiatric conditions, we introduced a form of sampling bias, specifically, selection bias, into the dataset. This bias could lead to misleading model performance if not properly accounted for during training.

To minimize this effect, we carefully curated the input features for the XGBoost models. Any features directly related to the traits excluded from the SuperControl group were removed from the training set. Without this step, the model might falsely infer that any psychiatric indicator implies the trait of interest, resulting in inflated, biologically uninformative predictions.

By excluding these confounding features, we aimed to prevent the model from learning trivial associations driven by the sampling design. Nonetheless, it’s important to emphasize that these models are not intended for clinical prediction or prospective diagnosis. Rather, they serve as reclassification tools, estimating the likelihood that individuals with partially overlapping psychiatric profiles may also be affected by the trait of interest.

This approach allows us to recover power lost due to sample exclusion while preserving trait-specific signals. However, the sampling bias inherent to the SuperControl definition means that imputed labels should be interpreted cautiously and only within the context of genetic association analyses.

### Genome wide association study (GWAS)

Genome-wide association analyses in FinnGen were conducted using REGENIE (v2.2.4)^44^ for all phenotypes. Models were adjusted for sex, birth year, age at death or end of follow-up (including a squared term), the first ten genetic principal components (PC1–10), and biobank-specific genotyping batch and chip effects (https://github.com/FINNGEN/regenie-pipelines). Quality control filters retained variants with minor allele frequency (MAF) > 0.001 and imputation info score > 0.8. Logistic regression was applied to all case-control traits.

### Conditional Analysis, Fine-Mapping, and Functional Annotation

Conditional association analyses were performed using GCTA (v1.94.1)^45^ to identify independent loci for the original FinnGen phenotypes, the Super Control phenotype, and the MTAG-derived phenotype. Fine-mapping of causal variants at each locus was conducted using the FinnGen fine-mapping pipeline, which integrates results from FINEMAP^46^ and SuSiE^47^. Fine-mapping regions were defined as ±1.5 Mb windows around each lead variant, consistent with recommendations for these tools.

### Genetic correlation, SNP-based heritability, and partitioned heritability

SNP-based heritability and genetic correlation were estimated using LD Score Regression (LDSC v1.0.1)^42^. Analyses were performed using the FinnGen “fin_ldsc” LD reference panel, a Finnish-specific LD resource designed to capture the unique genetic architecture of the Finnish population. To account for the binary nature of the GWAS traits and imbalanced case-control ratios, trait-specific effective sample sizes were calculated as:

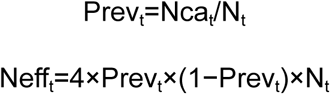

Where Prev is the prevalence, Nca is the number of cases, N is case plus control number and Neff is the effective sample size for each trait (t)

For MTAG-derived traits, effect sizes from MTAG output logs were used in the munging step. Summary statistics were processed for LDSC using the `munge_sumstats.py` script, with SNPs restricted to the HapMap 3 reference set. Because heritability estimates are influenced by trait prevalence in the cohort, direct comparison of observed heritability between the Original FinnGen and SuperControl Phenotypes is limited.

To further assess comparability, we considered liability-scale transformations to adjust for trait prevalence. However, this approach assumes appropriate case-control enrichment, which applies to the SuperControl Phenotype but not to the Original FinnGen Phenotype. Applying the transformation to the latter resulted in highly inflated heritability estimates, violating the model’s assumptions^48^. Therefore, all heritability estimates are reported on the observed scale.

Genetic correlations were computed using cross-trait LDSC (ldsc.py --rg). Partitioned heritability was assessed using the baseline v1.1 annotation model in combination with grouped cell-type specific annotations described by Finucane et al.^49^.

### GWAS exploration and replication evidence

To evaluate the novelty and reproducibility of genome-wide significant loci identified in the new phenotypes, we applied two complementary approaches: locus-based replication and gene-based replication.

In **locus-based replication (LBR)**, we compared lead SNPs and their surrounding ±1.5 Mb regions to previously published large-scale GWAS for each respective trait. For summary statistics not aligned to GRCh38, genomic coordinates were converted using the UCSC LiftOver tool^50^. A locus was classified as having **strong prior evidence** if a genome-wide significant variant (P < 5 × 10⁻⁸) was identified within the 3 Mb window in a prior GWAS. If no such variant was detected, but the lead SNP of the novel Phenotype reached P < 5 × 10⁻⁵ in a prior GWAS, the locus was labeled as having **moderate prior evidence**. Regions lacking any variant meeting these thresholds were considered to have **no supporting evidence**.

Studies used for this comparison were

Demontis 2023^51^ for ADHD,

Friligkou 2024^52^ for anxiety,

Mullins 2021^53^ and Connell 2025^54^ for bipolar disorder,

Howard 2019^55^ and Adams 2025^56^ for depression,

International Obsessive Compulsive Disorder Foundation (IOCDF) 2018^57^ and Strom 2025^58^ for OCD,

Nievergelt 2024^17^ for PTSD,

Trubetskoy 2022^18^ for Schizophrenia,

Watanabe 2022^59^ for sleep disorder and

And (CDG-PGC) 2019^60^ for overall psychiatric traits

For substance use disorder we included studies from different use disorders to cover all subtypes of this disorder. Therefore we included Sanchez-Roige 2019^61^, Walters 2018^62^, Saunders 2022^63^ for Alcohol use disorder, Johnson 2020^64^ for cannabis use disorder, Deak 2022^65^ and Polimanti 2020^66^ for opioid use disorder and Saunders 2022^63^ for smoking dependency.

In **gene-based replication (GBR)**, we evaluated whether mapped genes from each lead or fine-mapped variant had previously been associated with the same trait in the Open Targets Genetics platform^67^. Variant-to-gene (V2G) links incorporated functional annotations, including eQTLs, pQTLs, chromatin interactions, and in silico predictions. A variant was assigned **strong prior evidence** if at least one of its mapped genes was previously associated with the trait in Open Targets via GWAS. If the gene-trait link existed without supporting variant-level evidence, it was classified as having **moderate prior evidence**.

Variants without any gene- or variant-level support were considered to have **no supporting evidence**.

For comparison of LBR and GBR to account for the differing number of loci per trait, we evaluated the replication percentage per trait using the following metric:

Replication Percentage (RepPer) = (SPE + MPE) / (SPE + MPE + NSE)

where SPE = strong prior evidence, MPE = moderate prior evidence, and NSE = no supporting evidence. Statistical significance between the replication rates of the different Phenotypes was determined using the Wilcoxon Rank Sum test.

### Cross-cohort comparison

To evaluate the generalizability and cross-trait behavior of the Original, SuperControl, and PRISMA phenotypes, we performed cross-cohort prediction analyses in the UK Biobank (UKBB). Analyses were restricted to White European participants with available genetic principal components (PCs) (∼400,000 individuals).

#### Polygenic score computation and cross-cohort prediction

Polygenic risk scores (PGS) were generated for all phenotypes using the FinnGen PGS-weighting pipeline, which applies the PRS-CS algorithm for effect size shrinkage and SNP weight estimation. These PGS weights, derived from the FinnGen GWAS of the Original, SuperControl, and PRISMA phenotypes, were applied to UKBB genotype data to test cross-cohort predictability.

For each target trait, prediction models were fitted using a **generalized linear model (GLM)** of the form:

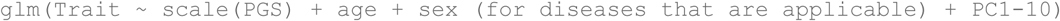

#### Regional polygenic scores based on non-replicated loci

To assess whether non-replicated loci contribute meaningful predictive signal across cohorts, we constructed regional PGS using SNP weights from genomic regions which were not replicated. Non-replicated regions were defined using LBR as 3 Mb windows (±1.5 Mb) surrounding lead variants. Comparison between phenotype definitions was performed using a two-sided z-test for the difference between independent log-odds ratios obtained from the Original, SuperControl, and PRISMA GWAS.

#### Ethics

Study subjects in FinnGen provided informed consent for biobank research, based on the Finnish Biobank Act. Alternatively, separate research cohorts, collected prior the Finnish Biobank Act came into effect (in September 2013) and start of FinnGen (August 2017), were collected based on study-specific consents and later transferred to the Finnish biobanks after approval by Fimea (Finnish Medicines Agency), the National Supervisory Authority for Welfare and Health. Recruitment protocols followed the biobank protocols approved by Fimea. The Coordinating Ethics Committee of the Hospital District of Helsinki and Uusimaa (HUS) statement number for the FinnGen study is Nr HUS/990/2017.

The FinnGen study is approved by Finnish Institute for Health and Welfare (permit numbers: THL/2031/6.02.00/2017, THL/1101/5.05.00/2017, THL/341/6.02.00/2018, THL/2222/6.02.00/2018, THL/283/6.02.00/2019, THL/1721/5.05.00/2019 and THL/1524/5.05.00/2020), Digital and population data service agency (permit numbers: VRK43431/2017-3, VRK/6909/2018-3, VRK/4415/2019-3), the Social Insurance Institution (permit numbers: KELA 58/522/2017, KELA 131/522/2018, KELA 70/522/2019, KELA 98/522/2019, KELA 134/522/2019, KELA 138/522/2019, KELA 2/522/2020, KELA 16/522/2020), Findata permit numbers THL/2364/14.02/2020, THL/4055/14.06.00/2020, THL/3433/14.06.00/2020, THL/4432/14.06/2020, THL/5189/14.06/2020, THL/5894/14.06.00/2020, THL/6619/14.06.00/2020, THL/209/14.06.00/2021, THL/688/14.06.00/2021, THL/1284/14.06.00/2021, THL/1965/14.06.00/2021, THL/5546/14.02.00/2020, THL/2658/14.06.00/2021, THL/4235/14.06.00/2021, THL/4990/14.02.00/2023 Statistics Finland (permit numbers: TK-53-1041-17 and TK/143/07.03.00/2020 (earlier TK-53-90-20) TK/1735/07.03.00/2021, TK/3112/07.03.00/2021) and Finnish Registry for Kidney Diseases permission/extract from the meeting minutes on 4th July 2019.

The Biobank Access Decisions for FinnGen samples and data utilized in FinnGen Data Freeze 13 include: THL Biobank BB2017_55, BB2017_111, BB2018_19, BB_2018_34, BB_2018_67, BB2018_71, BB2019_7, BB2019_8, BB2019_26, BB2020_1, BB2021_65, BB22-0025-A01, BB22-0025-A03, BB23-0222-A01, BB22-0025-A04, BB22-0025-A06, BB22-0025-A08, THLBB2024_30. Finnish Red Cross Blood Service Biobank 7.12.2017, 13.11.2023, 001-2023, Helsinki Biobank HUS/359/2017, HUS/248/2020, HUS/430/2021 §28, §29, HUS/150/2022 §12, §13, §14, §15, §16, §17, §18, §23, §58, §59, HUS/128/2023 §18, BB22-0025-A01, BB22-0025-A02, BB22-0025-A05, BB22-0025-A07, BB22-0025-A09, BB22-0025-A10, BB22-0025-A03, BB23-0222-A01, BB22-0025-A04, BB22-0025-A06, BB22-0025-A08, Amendment_BB22-0025-A05, Decision allowing to continue data processing until 31st Aug 2027: BB_2021-0140, HUS/150/2022 §12, BB_2021-0139, HUS/150/2022 §13, BB_2021-0161,HUS/150/2022 §14, BB_2021-0164, HUS/150/2022 §15, BB_2021-0169, HUS/150/2022 §16, BB_2021-0170, HUS/150/2022 §17, BB_2021-0179, HUS/150/2022 §18, BB_2022-0262, HUS/150/2022 §58, BB22-0067, HUS/150/2022 §59, Auria Biobank AB17-5154 and amendment #1 (August 17 2020) and amendments BB_2021-0140, BB_2021-0156 (August 26 2021, Feb 2 2022), BB_2021-0169, BB_2021-0179, BB_2021-0161, AB20-5926 and amendment #1 (April 23 2020) and it’s modifications (Sep 22 2021), BB_2022-0262, BB_2022-0256, BB22-0025-A01, BB22-0025-A02, BB22-0025-A03, BB23-0222_A01, BB22-0025-A02, BB22-0025-A05, BB22-0025-A07, BB22-0025-A09, BB22-0025-A10, BB22-0025-A03, BB23-0222-A01, BB22-0025-A04, BB22-0025-A06, BB22-0025-A08, Decision allowing to continue data processing until 31st Aug 2027: AB20-5926, BB_2021- 0140, BB_2021-0156, BB_2021-0161, BB_2021- 0161, BB_2021-0164, BB_2021-0169, BB_2021-0179, BB_2022-0262, Biobank Borealis of Northern Finland_2017_1013, 2021_5010, 2021_5010 Amendment, 2021_5018, 2021_5018 Amendment, 2021_5015, 2021_5015 Amendment, 2021_5015 Amendment_2, 2021_5023, 2021_5023 Amendment, 2021_5023 Amendment_2, 2021_5017, 2021_5017 Amendment, 2022_6001, 2022_6001 Amendment, 2022_6006 Amendment, 2022_6006 Amendment_2, BB22-0067, 2022_0262, 2022_0262 Amendment, BB22-0025-A01, BB22-0025-A02, BB22-0025-A05, BB22-0025-A07, BB22-0025-A09, BB22-0025-A10, BB22-0025-A03, BB23-0222-A01, BB22-0025-A04, BB22-0025-A06, BB22-0025-A08, Decision allowing to continue data processing until 31st Aug 2027: BB/2021/5015, BB/2021/5017, BB/2021/5018, BB/2021/5023, BB/2022/6006, BB/2022/6001, BB/2022-0262, BB/2021/5010, Biobank of Eastern Finland 1186/2018 and amendment 22§/2020, 53§/2021, 13§/2022, 14§/2022, 15§/2022, 27§/2022, 28§/2022, 29§/2022, 33§/2022, 35§/2022, 36§/2022, 37§/2022, 39§/2022, 7§/2023, 32§/2023, 33§/2023, 34§/2023, 35§/2023, 36§/2023, 37§/2023, 38§/2023, 39§/2023, 40§/2023, 41§/2023, BB22-0025-A01, BB22-0025-A02, BB22-0025-A05, BB22-0025-A07, BB22-0025-A09, BB22-0025-A10, BB22-0025-A03, BB23-0222-A01, BB22-0025-A04, BB22-0025-A06, BB22-0025-A08, Decision allowing to continue data processing until 31st Aug 2027: MO-BB_2021-0179-A0, MO-BB_2021-0156_PRE-A01, BB_2021-0140, MO-BB_2021-0170_PRE-A0, MO-BB_2021-0169-A01, MO-BB_2022-0256-A01, MO-BB_2021-0161-A01, MO-BB_2021-0161-A02, BB22-0067-A01, MO-BB_2022-0262-A0, Finnish Clinical Biobank Tampere MH0004 and amendments (21.02.2020 & 06.10.2020), BB2021-0140 8§/2021, 9§/2021, §9/2022, §10/2022, §12/2022, 13§/2022, §20/2022, §21/2022, §22/2022, §23/2022, 28§/2022, 29§/2022, 30§/2022, 31§/2022, 32§/2022, 38§/2022, 40§/2022, 42§/2022, 1§/2023, BB2021-0140, BB22-0025-A01, BB_2021-0161, BB22-0025-A02, BB22-0025-A05, BB22-0025-A07, BB22-0025-A09, BB22-0025-A10, BB22-0025-A03, BB23-0222-A01, BB22-0025-A04, BB22-0025-A06, BB22-0025-A08, Decision allowing to continue data processing until 31st Aug 2027: BB_2021- 0140, BB_ 2021- 0161, BB_ 2021- 0179, BB_ 2021- 0156, BB_ 2021- 0169, BB_ 2021- 0170, BB22- 0067- A01, Central Finland Biobank 1-2017, BB_2021-0169, BB_2021-0179, BB_2022-0256, BB_2022-0262, Decision allowing to continue data processing until 31st Aug 2027 for projects: BB_2021-0179, BB22-0067,BB_2022-0262, BB_2021-0170, BB_2021-0164, BB_2021-0161, and BB_2021-0169, BB22-0025-A01, BB22-0025-A02, BB22-0025-A05, BB22-0025-A07, BB22-0025-A09, BB22-0025-A10, BB22-0025-A03, BB23-0222-A01, BB22-0025-A04, BB22-0025-A06, BB22-0025-A08, Terveystalo Biobank STB 2018001 and amendment 25th Aug 2020, Finnish Hematological Registry and Clinical Biobank decision 18th June 2021, Amendment 2nd January 2024 and Arctic biobank P0844: ARC_2021_1001, ARC_2023_3003 (BB22-0025-A01), BB22-0025-A03, BB23-0222-A01, BB22-0025-A04, BB22-0025-A06, BB22-0025-A08.

## Data Availability

All sharable data produced in the present study are available upon reasonable request to the authors

## Acknowledgements

The authors thank Anniina Tervi for her helpful feedback and advice during the revision process. We also gratefully acknowledge all participants of the FinnGen project for their invaluable contributions to this research.

The author gratefully acknowledges funding support from the University of Helsinki Research Foundation and the Vilho, Yrjö ja Kalle Väisälän rahasto of the Finnish Academy of Science and Letters, which supported the doctoral studies underlying this work

