## Supplement-Material for "Reducing misclassification bias in electronic health record-based GWAS of psychiatric traits using the SuperControl framework"

### FinnGen Study Cohort and Genetic Data

#### Cohort Overview

FinnGen is a nationwide research project that integrates genotype data from over 520,000 Finnish biobank participants with longitudinal national health register data. The cohort includes individuals recruited through hospital biobanks, disease-specific studies, and blood donor registries, with intentional enrichment for disease cases to enhance statistical power for genetic discovery. FinnGen participants had a median age of 53 years at sample collection, with 57% female and 43% male.

#### Genotyping and Quality Control

Participants were genotyped using custom ThermoFisher Axiom arrays: 121,000 samples on version 1 (657,675 markers) and ~328,000 on version 2 (723,376 probe sets for 664,510 markers). Arrays include GWAS backbones and variants relevant to coding regions, pharmacogenomics, immune traits, and Finnish-enriched alleles.

Sample-level QC excluded individuals based on:

- Sex discordance between genetically inferred and registry-reported sex (F-statistic $< 0.4$ for females, $> 0.7$ for males),
- Genotype missingness $> 2\%$,
- Excess heterozygosity in common variants (allele frequency $> 0.05$, $> 3$ SD from batch mean),
- Excess relatedness ($\hat{\pi} > 0.1$), removed in two rounds: first for samples related to $> 500$ others, then for those related to $> 50$ others.

Variant-level QC removed markers that:

- Had non-ATCG alleles,
- Were absent from the SISu v4.2 imputation panel or had panel allele frequency $< 0.001$ (imputation only),
- Showed significant allele frequency deviation from the panel ($p < 5\times10^{-8}$, adjusted for 10 PCs),
- Had Hardy-Weinberg disequilibrium p-value $< 1\times10^{-10}$ across all batches (with exceptions for rare variants with homozygote deficiency),
- Were missing in $> 15\%$ of batches or non-PASS in $> 30\%$ of batches.

#### Imputation and Ancestry Assignment

Genotyped samples were pre-phased with Eagle 2.3.5 using 20,000 conditioning haplotypes and imputed using the Finnish-specific SISu v4.2 reference panel. The panel was built from high-coverage (25x) whole-genome sequencing of 8,554 Finnish individuals and mapped to GRCh38.

Ancestry was implicitly accounted for by using a Finnish-specific reference panel, eliminating the need for global ancestry prediction. Population outliers and samples with excessive relatedness were removed as part of quality control to reduce cryptic structure.

#### Phenotype Definition from Health Registers

FinnGen uses harmonized clinical phenotype definitions based on structured data from a wide range of nationwide health and social care registers. The following data sources were used:

- Statistics Finland
- Finnish Cancer Registry and Mass Screening Registry
- Register of Primary Health Care Visits (Avohilmo)
- Care Register for Health Care
- The Social Insurance Institution of Finland (Kela)
- Digital and Population Data Services Agency
- Finnish Register of Visual Impairment
- Care Register for Social Welfare
- Finnish Registry for Kidney Diseases
- Finnish National Infectious Diseases Register
- Medical Birth Register
- Finnish National Vaccination Register
- Register of Congenital Malformations

These registers provide detailed information on diagnoses, procedures, prescriptions, hospitalizations, and mortality. Phenotypes were defined using harmonized algorithms developed by expert working groups and are publicly available. Socioeconomic covariates were based on Statistics Finland's classification of education levels (https://stat.fi/fi/luokitukset/koulutusaste/koulutusaste_1_20160101). Occupational categories were assigned using the national classification of occupations (https://stat.fi/en/luokitukset/ammatti/ammatti_1_20010101). This study used definitions from FinnGen Data Freeze 12 (DF12).

### UK Biobank Study Cohort and Genetic Data

**Cohort Overview**
UK Biobank is a population-based prospective cohort of approximately 500,000 individuals aged 40–69 at recruitment between 2006 and 2010. Participants provided biological samples and consent for long-term follow-up via linkage to national health records.

**Genotyping and Quality Control**
Participants were genotyped using two closely related arrays: 49,950 individuals with the UK BiLEVE Axiom array and ~438,000 with the UK Biobank Axiom array. Both arrays share >95% marker content and include ~800,000 variants selected for genome-wide imputation coverage and trait relevance. Genotyping was performed at the Affymetrix Research Services Laboratory in batches of ~4,800 samples.

Variant-level QC was applied batch-wise using Affymetrix protocols and custom filters. Variants were excluded if they exhibited:

- Hardy-Weinberg disequilibrium ($p < 1\times10^{-12}$),
- Plate or batch effects ($p < 1\times10^{-12}$ via Fisher's exact test),
- 2% missingness across samples,
- Poor clustering or discordance in replicate control samples.

Samples were excluded for:

- Sex mismatches (self-reported vs. genetic),

- Excess heterozygosity or missingness (adjusted for population structure),
- Sex chromosome aneuploidy,
- Relatedness or duplicates, if unresolved.

**Imputation and Ancestry Assignment**
Imputation was performed for 487,442 individuals using IMPUTE4 with a combined reference panel from the Haplotype Reference Consortium (HRC) and UK10K + 1000 Genomes. Variants were retained only if present in both panels, resulting in ~97 million imputed SNPs.

Ancestry inference used PCA on a curated set of ~467,000 autosomal SNPs shared with HapMap3 and present in the imputed data. Individuals were assigned to five super-populations (AFR, AMR, EAS, EUR, SAS) using GenoPred's multinomial elastic-net classifier trained on 1000 Genomes data. Outliers were excluded based on Mahalanobis distance from ancestry-specific centroids.

**Phenotype Definition from Health Registers**
UK Biobank provides linked health data including hospital inpatient diagnoses (ICD-9/10, OPCS-3/4), death records (ICD-10), primary care data (READ2, CTV-3), and cancer registry entries. Phenotypes were derived using first occurrence mappings and algorithmically-defined outcomes curated from structured health records. Censoring dates for hospital, cancer, and death data were set at 31 January 2021.

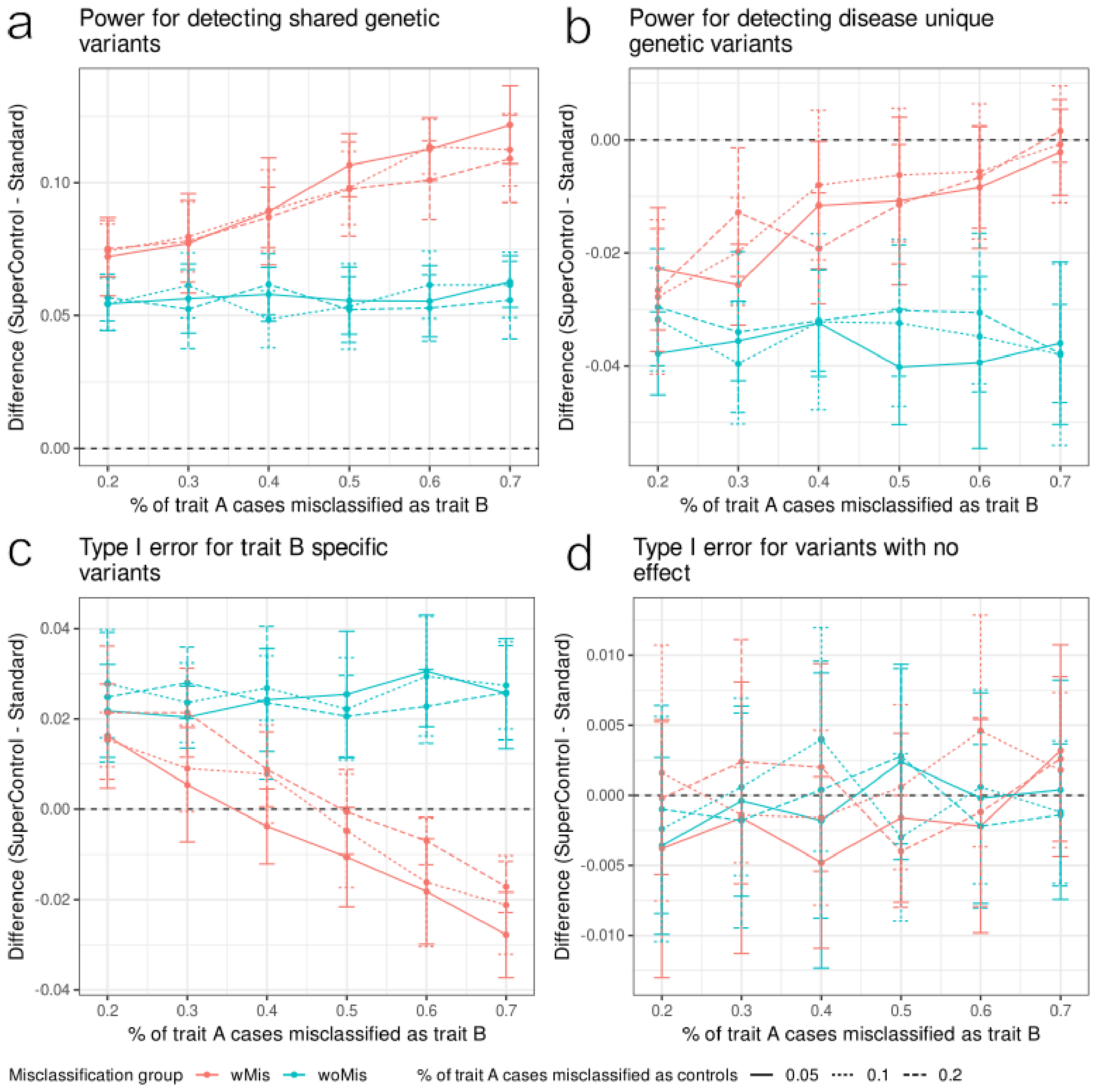


**Supplementary Fig. 1 | Simulation of SuperControl and Standard phenotypes under varying misclassification at 30% trait prevalence.**

a-d, Differences (SuperControl – Standard) in power or type I error across increasing levels of misclassification in trait B and among controls other than trait B. a, Power for SNPs shared between traits A and B. b, Power for SNPs specific to trait A. c,Type I error for SNPs causal only for trait B. d, Type I error for non-causal SNPs. Positive values indicate higher numerical values under the SuperControl definition, and negative values indicate higher values under the Standard definition. Curves represent the mean of ten independent simulations, with error bars denoting standard deviation.

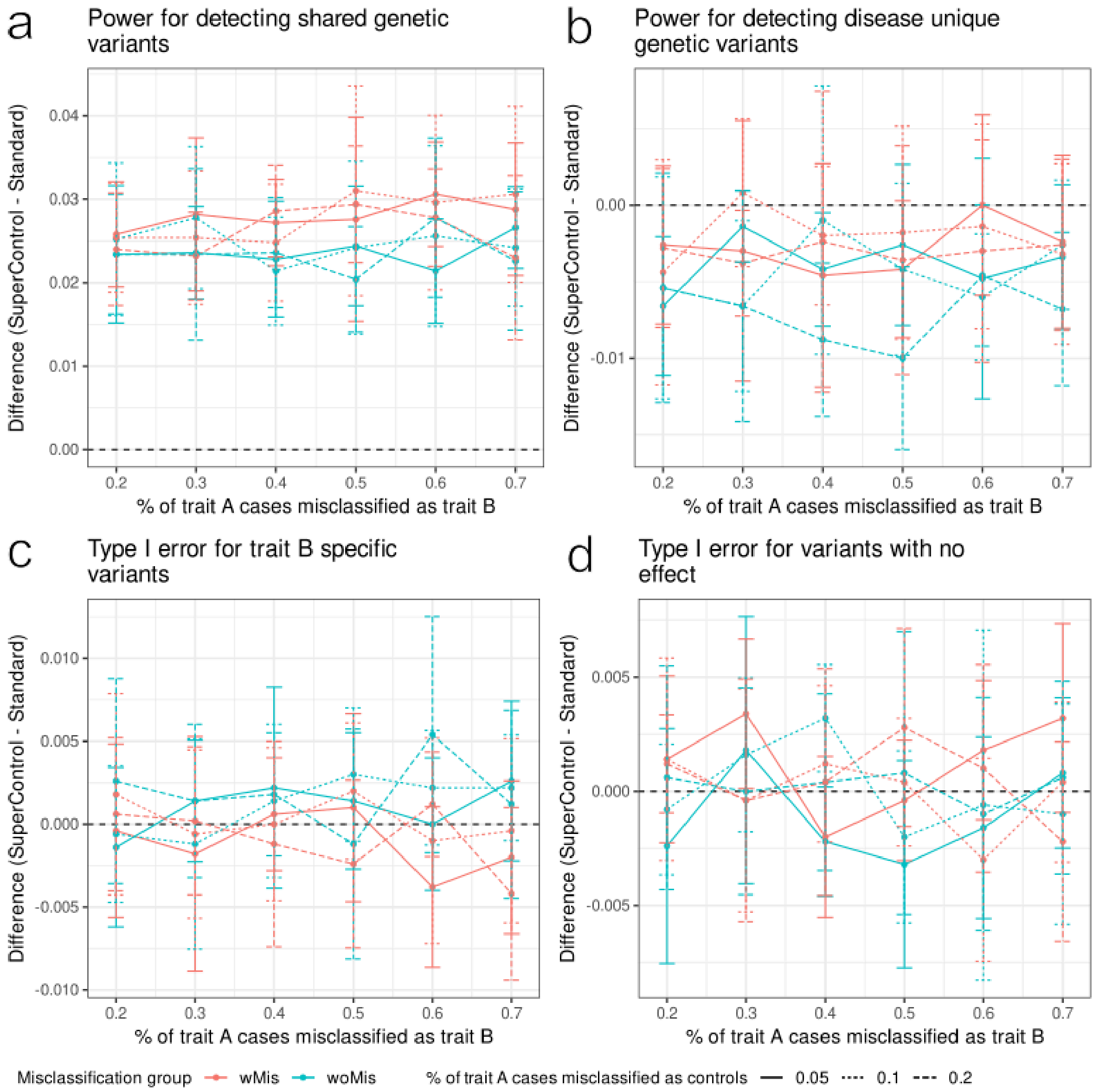


**Supplementary Fig. 2 | Simulation of SuperControl and Standard phenotypes under varying misclassification at 10% trait prevalence.**

a-d, Differences (SuperControl – Standard) in power or type I error across increasing levels of misclassification in trait B and among controls other than trait B. a, Power for SNPs shared between traits A and B. b, Power for SNPs specific to trait A. c,Type I error for SNPs causal only for trait B. d, Type I error for non-causal SNPs. Positive values indicate higher numerical values under the SuperControl definition, and negative values indicate higher values under the Standard definition. Curves represent the mean of ten independent simulations, with error bars denoting standard deviation.

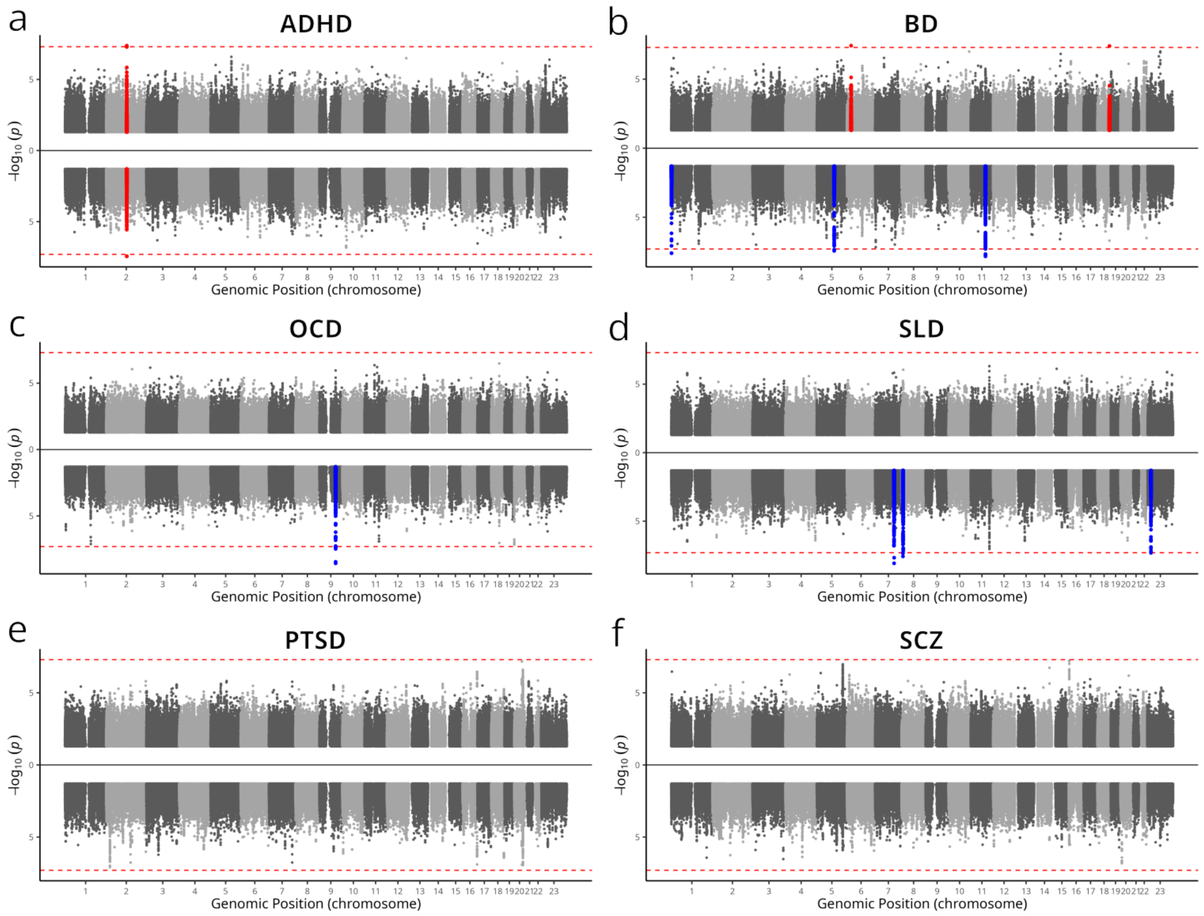


**Supplementary Fig. 3 | Manhattan plot of Genome-wide significant loci in Original FinnGen and SuperControl phenotypes across psychiatric traits.**

a-f, Mirrored Manhattan plots for attention-deficit/hyperactivity disorder (ADHD), bipolar disorder (BD), obsessive-compulsive disorder (OCD), sleep disorders (SLD), post-traumatic stress disorder (PTSD) and schizophrenia (SCZ). In each plot, the upper half represents results from the Original FinnGen phenotype and the lower half from the SuperControl phenotype. Red points denote loci shared between the two phenotypes or unique to the Original FinnGen phenotype, and blue points denote loci unique to the SuperControl phenotype.

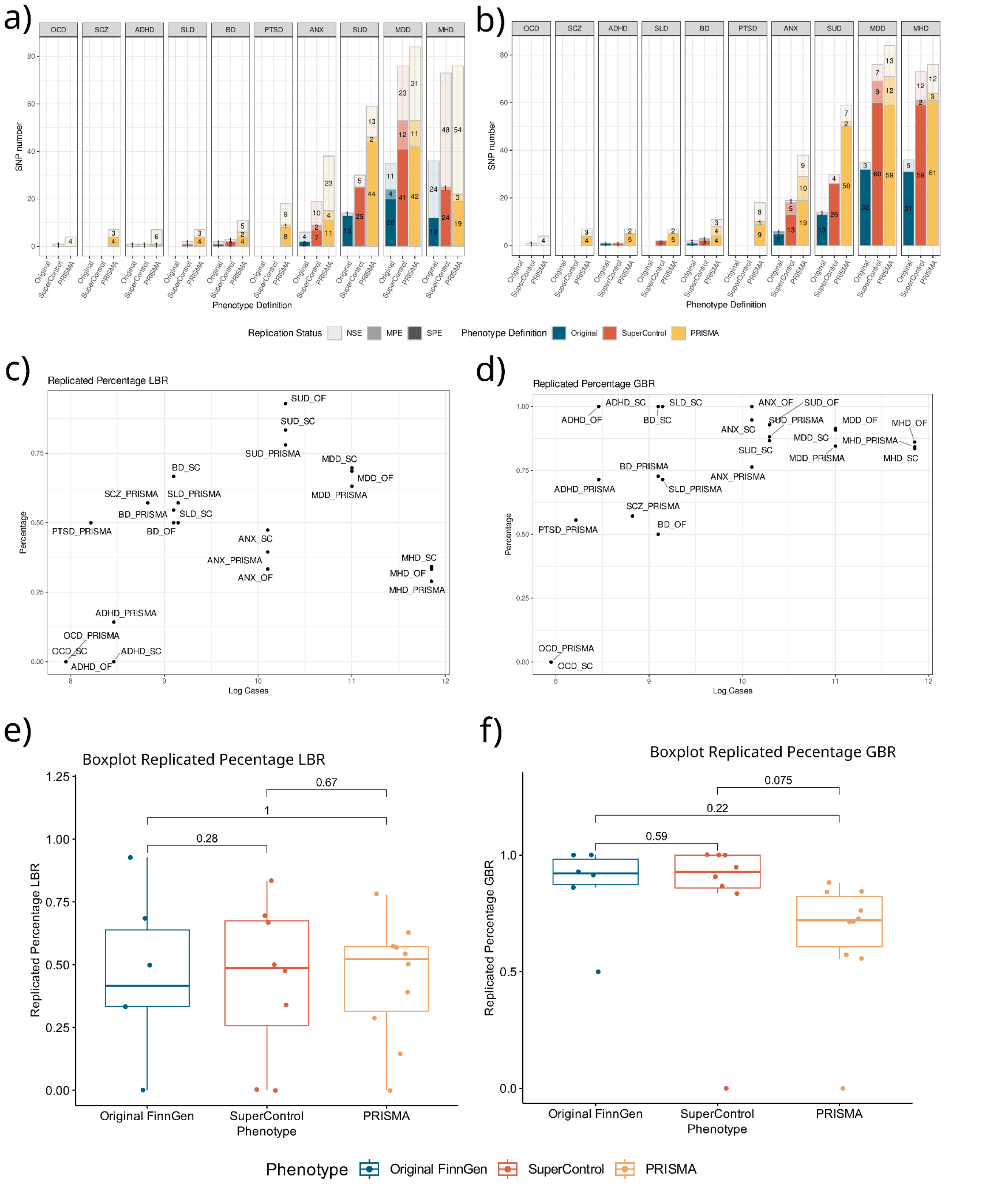

**Supplementary Fig. 4 | Replication of independent loci in Original FinnGen, SuperControl, and PRISMA phenotypes across psychiatric traits.**
a-b, Barplots showing locus-based replication (LBR, a) and gene-based replication (GBR, b) for ten psychiatric traits: SCZ, MDD, BD, ANX, ADHD, PTSD, OCD, SLD, SUD, and the combined “any mental health disorder” trait (MHD). Each trait contains three bars representing loci unique to the Original FinnGen phenotype (left), the SuperControl phenotype (middle), and the PRISMA phenotype (right). Bars are segmented by replication strength: strong prior evidence (SPE, darkest shading), moderate prior evidence (MPE, intermediate shading), and no supporting evidence (NSE, faintest shading). c-d, Replication rates for LBR (c) and GBR (d), defined as the percentage of loci or genes with either SPE or MPE among all loci or genes detected. Results are shown for all ten traits across the three phenotypes. e-f, Boxplots summarizing replication rates across all ten traits for each phenotype, shown separately for LBR (e) and GBR (f). Pairwise differences in replication rates between phenotypes were tested using the Wilcoxon rank-sum test.

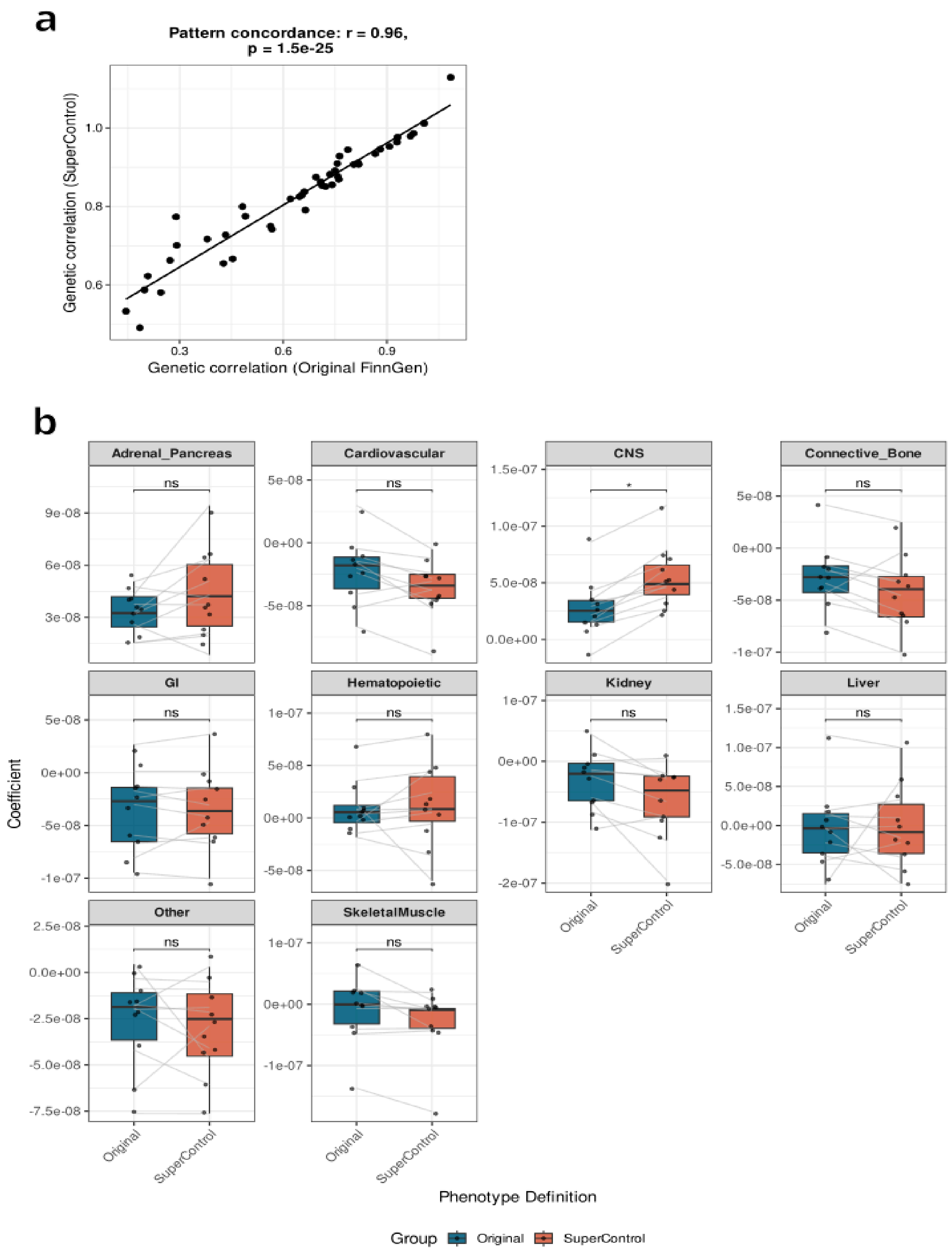


**Figure 5 | Comparison of genetic correlations between Original and SuperControl phenotypes.** A, Scatter plot showing pairwise genetic correlations across ten psychiatric traits estimated using LDSC for the Original FinnGen and SuperControl definitions. Each point represents one trait pair. The overall correlation between definitions was high (Pearson $r = 0.96$, $p < 2.2 \times 10^{-16}$), demonstrating preserved genetic structure. Mean correlations were moderately higher under the SuperControl definition ($\Delta = +0.19$, $p = 6.5 \times 10^{-13}$), reflecting a small but systematic increase in shared signal. B, Boxplot of Coefficient values of partitioned heritability by tissue. Paired Wilcoxon rank sum test to compare effect sizes between the original FinnGen Phenotype (blue) and the SuperControl Phenotype (red)

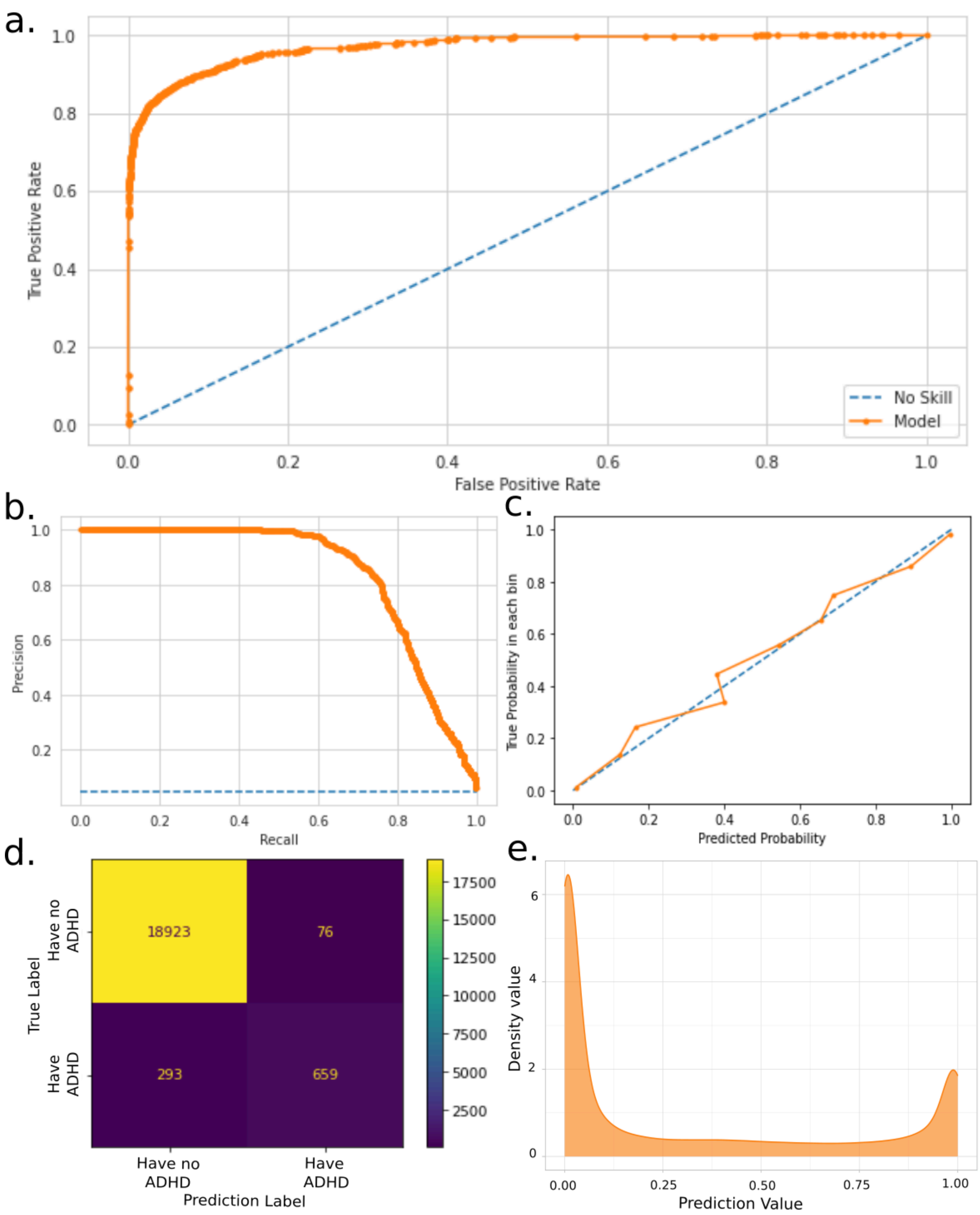


**Supplementary Fig. 6 | Evaluation of XGBoost model performance in predicting ADHD.**

Performance metrics for XGBoost classifier trained to distinguish ADHD cases from individuals with no recorded mental health diagnoses (super control definition). Model evaluation was conducted on a held-out validation set comprising 20% of the data. a) Receiver operating characteristic (ROC) curve (AUC = 98.15%). b) Precision-Recall curve. c) Calibration curve for predicted probabilities and observed outcomes. d) Confusion matrix displaying classification results on the validation set. e) Density plot showing predicted probabilities for those excluded from the super control phenotype.

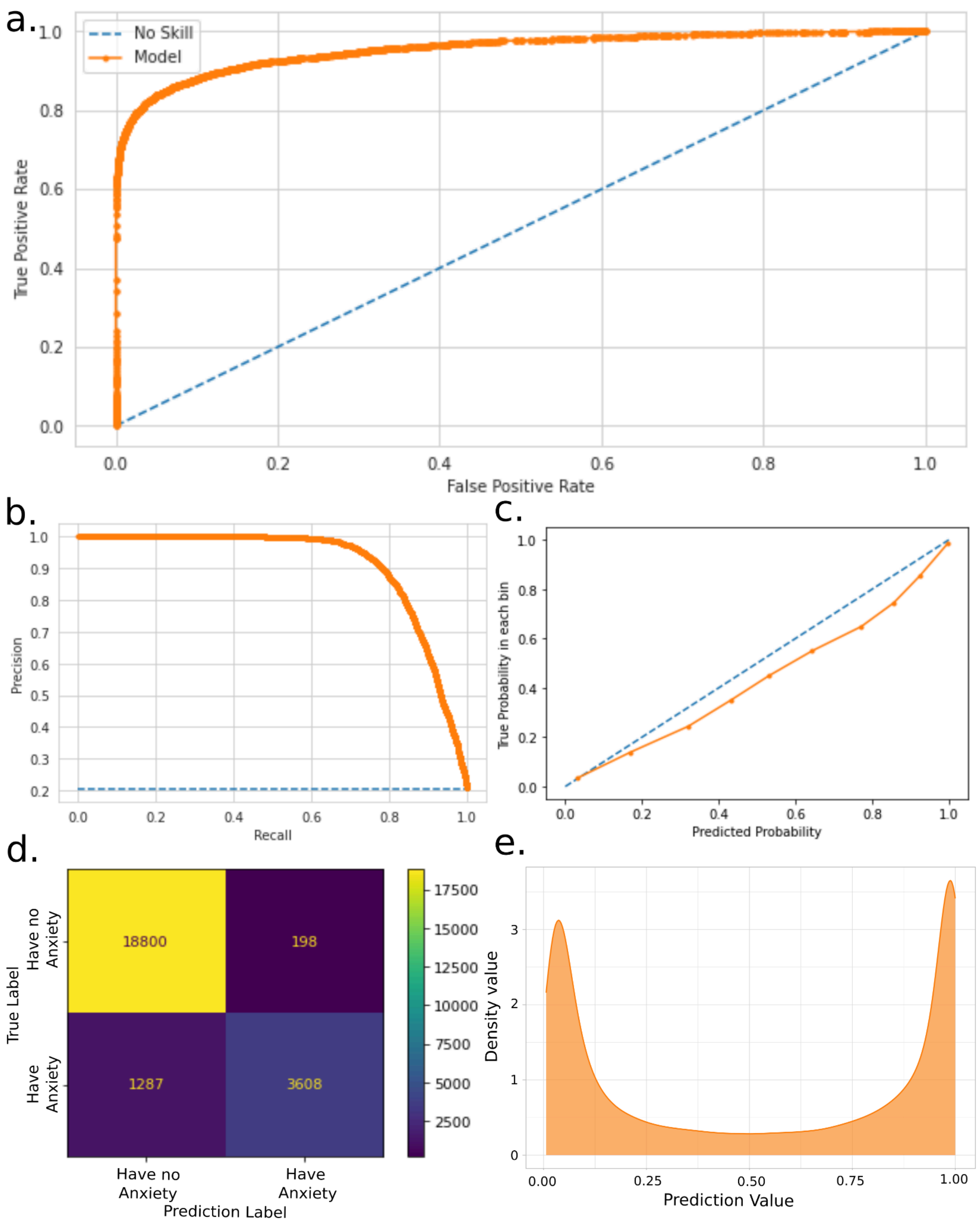


**Supplementary Fig. 7 | Evaluation of XGBoost model performance in predicting Anxiety.**

Performance metrics for XGBoost classifier trained to distinguish Anxiety cases from individuals with no recorded mental health diagnoses (super control definition). Model evaluation was conducted on a held-out validation set comprising 20% of the data. a) Receiver operating characteristic (ROC) curve (AUC = 93.78%). b) Precision-Recall curve. c) Calibration curve for predicted probabilities and observed outcomes. d) Confusion matrix displaying classification results on the validation set. e) Density plot showing predicted probabilities for those excluded from the super control phenotype.

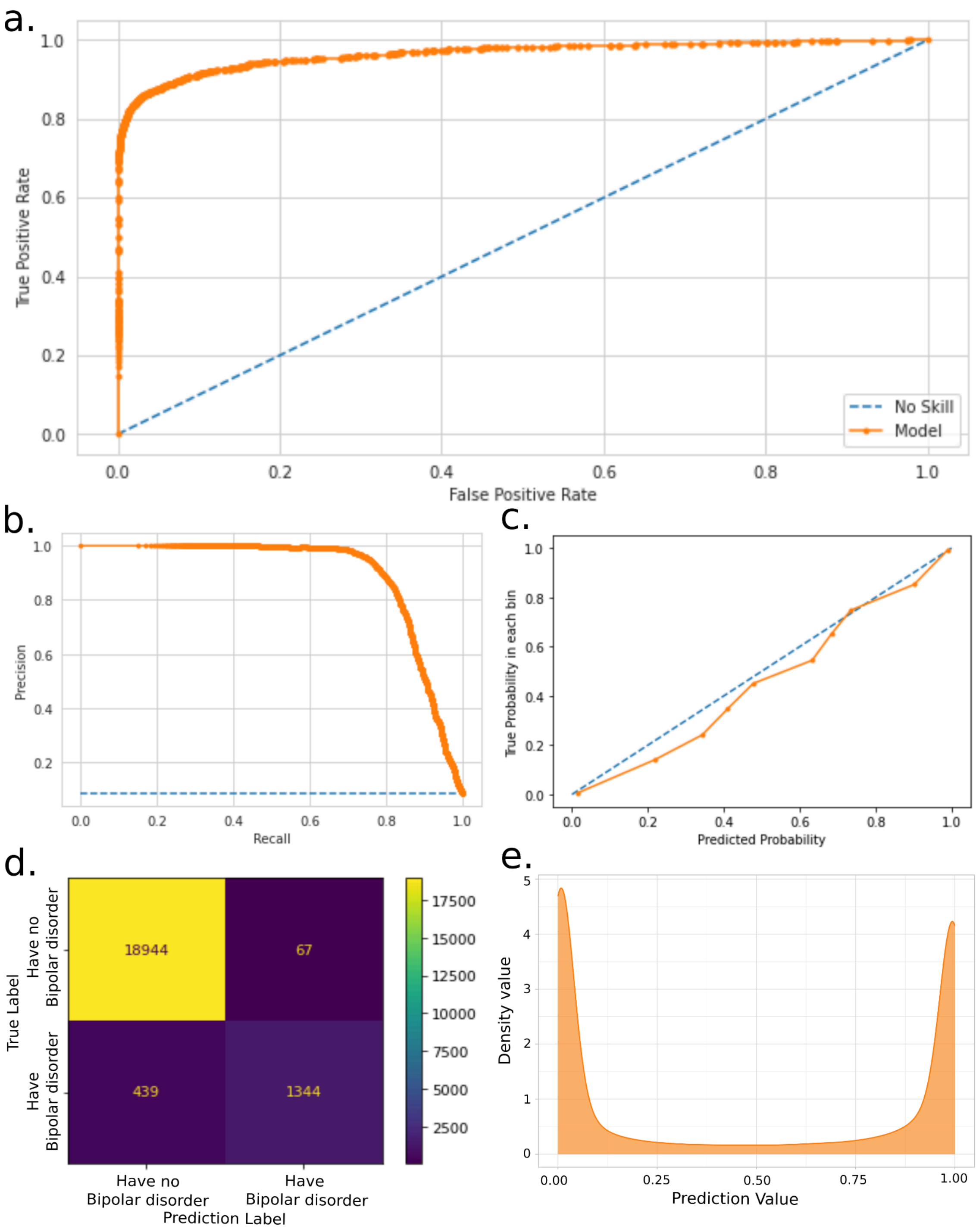


**Supplementary Fig. 8 | Evaluation of XGBoost model performance in predicting Bipolar disorder.**

Performance metrics for XGBoost classifier trained to distinguish Bipolar disorder cases from individuals with no recorded mental health diagnoses (super control definition). Model evaluation was conducted on a held-out validation set comprising 20% of the data. a) Receiver operating characteristic (ROC) curve (AUC = 97.57%). b) Precision-Recall curve. c) Calibration curve for predicted probabilities and observed outcomes. d) Confusion matrix displaying classification results on the validation set. e) Density plot showing predicted probabilities for those excluded from the super control phenotype.

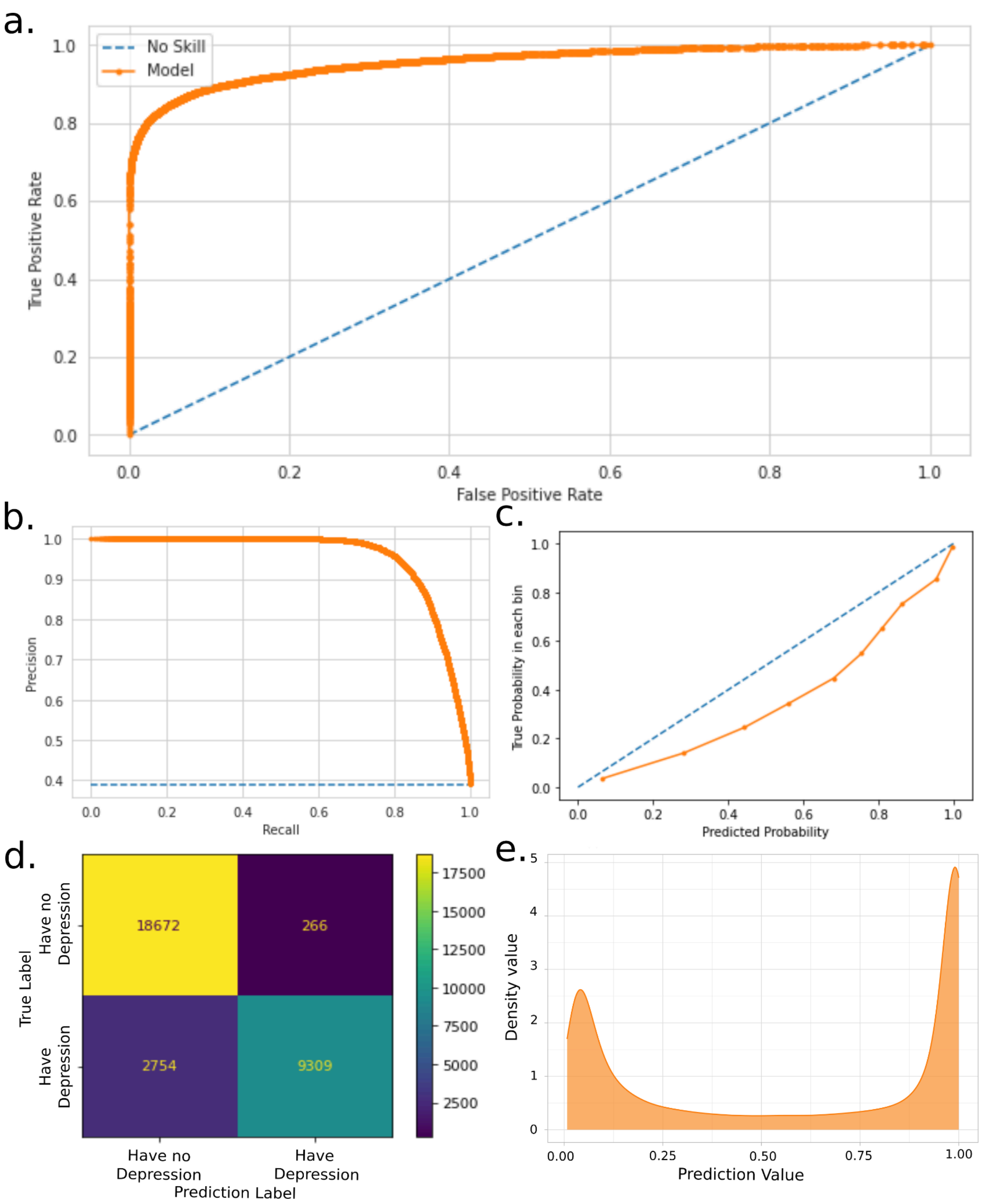


**Supplementary Fig. 9 | Evaluation of XGBoost model performance in predicting Depression.**

Performance metrics for XGBoost classifier trained to distinguish Depression cases from individuals with no recorded mental health diagnoses (super control definition). Model evaluation was conducted on a held-out validation set comprising 20% of the data. a) Receiver operating characteristic (ROC) curve (AUC = 90.26%). b) Precision-Recall curve. c) Calibration curve for predicted probabilities and observed outcomes. d) Confusion matrix displaying classification results on the validation set. e) Density plot showing predicted probabilities for those excluded from the super control phenotype.

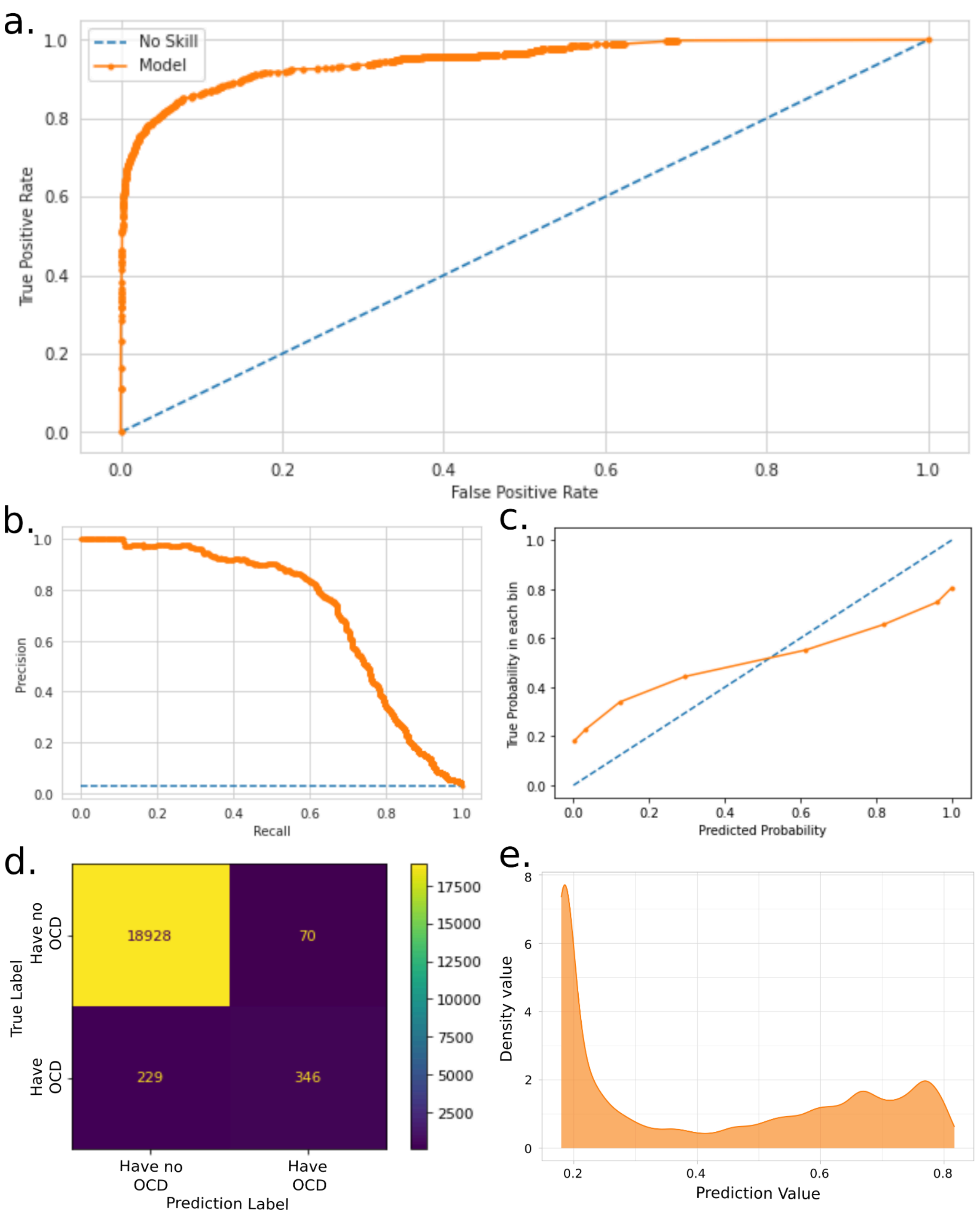


**Supplementary Fig. 10 | Evaluation of XGBoost model performance in predicting OCD.**

Performance metrics for XGBoost classifier trained to distinguish OCD cases from individuals with no recorded mental health diagnoses (super control definition). Model evaluation was conducted on a held-out validation set comprising 20% of the data. a) Receiver operating characteristic (ROC) curve (AUC = 98.47%). b) Precision-Recall curve. c) Calibration curve for predicted probabilities and observed outcomes. d) Confusion matrix displaying classification results on the validation set. e) Density plot showing predicted probabilities for those excluded from the super control phenotype.

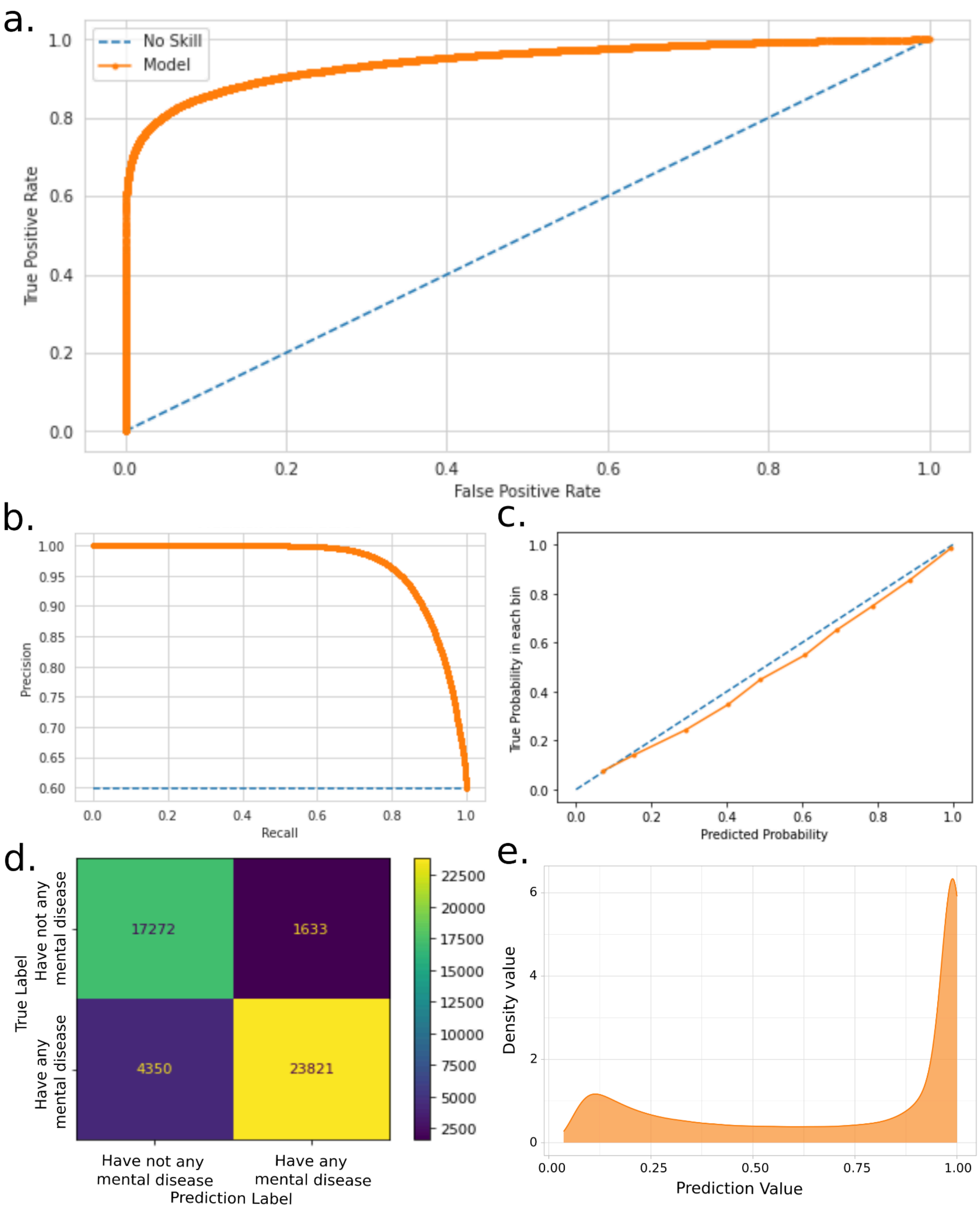


**Supplementary Fig. 11 | Evaluation of XGBoost model performance in predicting mental diseases.**

Performance metrics for XGBoost classifier trained to distinguish mental diseases cases from individuals with no recorded mental health diagnoses (super control definition). Model evaluation was conducted on a held-out validation set comprising 20% of the data. a) Receiver operating characteristic (ROC) curve (AUC = 87.29%). b) Precision-Recall curve. c) Calibration curve for predicted probabilities and observed outcomes. d) Confusion matrix displaying classification results on the validation set. e) Density plot showing predicted probabilities for those excluded from the super control phenotype.

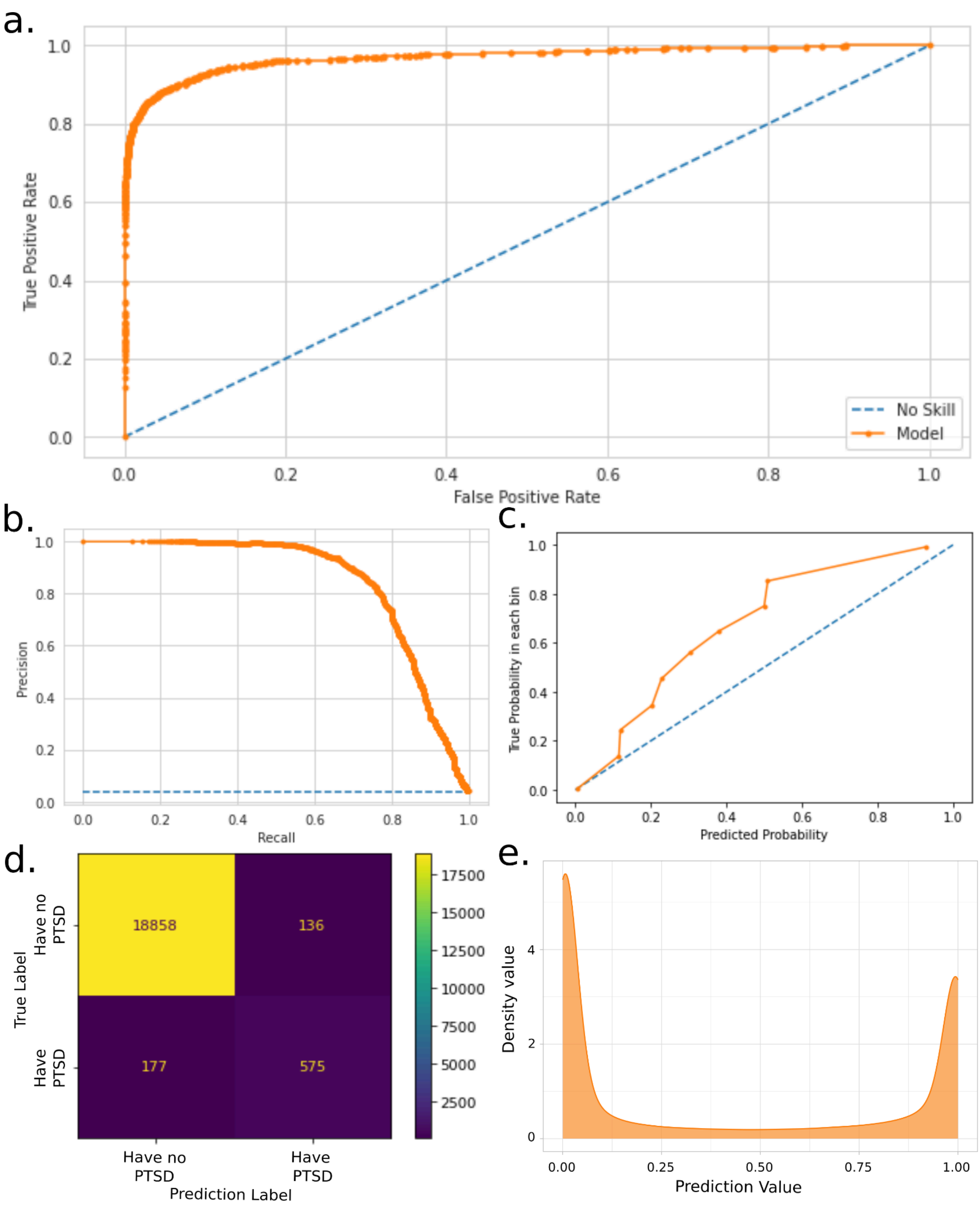


**Supplementary Fig. 12 | Evaluation of XGBoost model performance in predicting PTSD.**

Performance metrics for XGBoost classifier trained to distinguish PTSD cases from individuals with no recorded mental health diagnoses (super control definition). Model evaluation was conducted on a held-out validation set comprising 20% of the data. a) Receiver operating characteristic (ROC) curve (AUC = 98.41%). b) Precision-Recall curve. c) Calibration curve for predicted probabilities and observed outcomes. d) Confusion matrix displaying classification results on the validation set. e) Density plot showing predicted probabilities for those excluded from the super control phenotype.

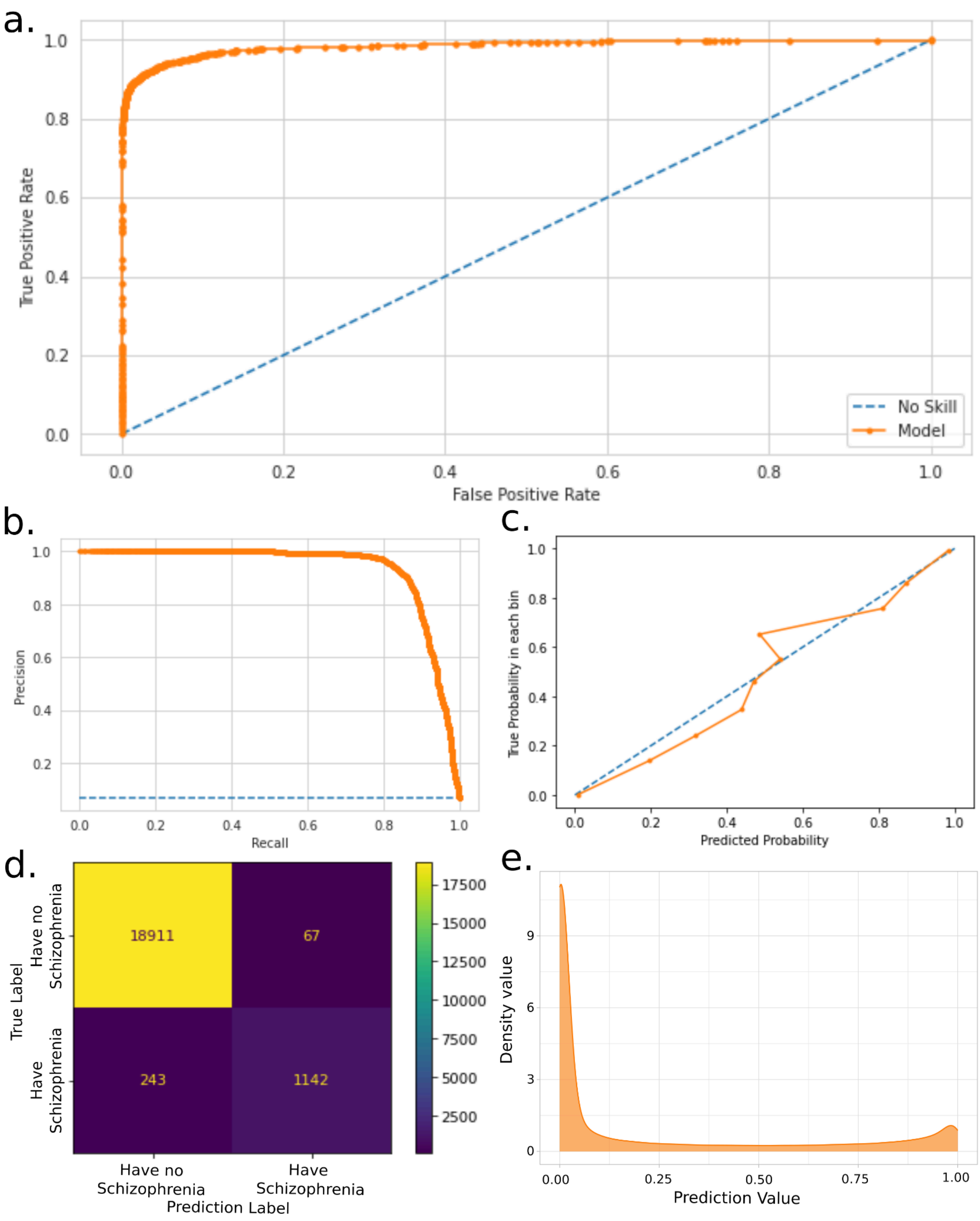


**Supplementary Fig. 13 | Evaluation of XGBoost model performance in predicting Schizophrenia.**

Performance metrics for XGBoost classifier trained to distinguish Schizophrenia cases from individuals with no recorded mental health diagnoses (super control definition). Model evaluation was conducted on a held-out validation set comprising 20% of the data. a) Receiver operating characteristic (ROC) curve (AUC = 98.48%). b) Precision-Recall curve. c) Calibration curve for predicted probabilities and observed outcomes. d) Confusion matrix displaying classification results on the validation set. e) Density plot showing predicted probabilities for those excluded from the super control phenotype.

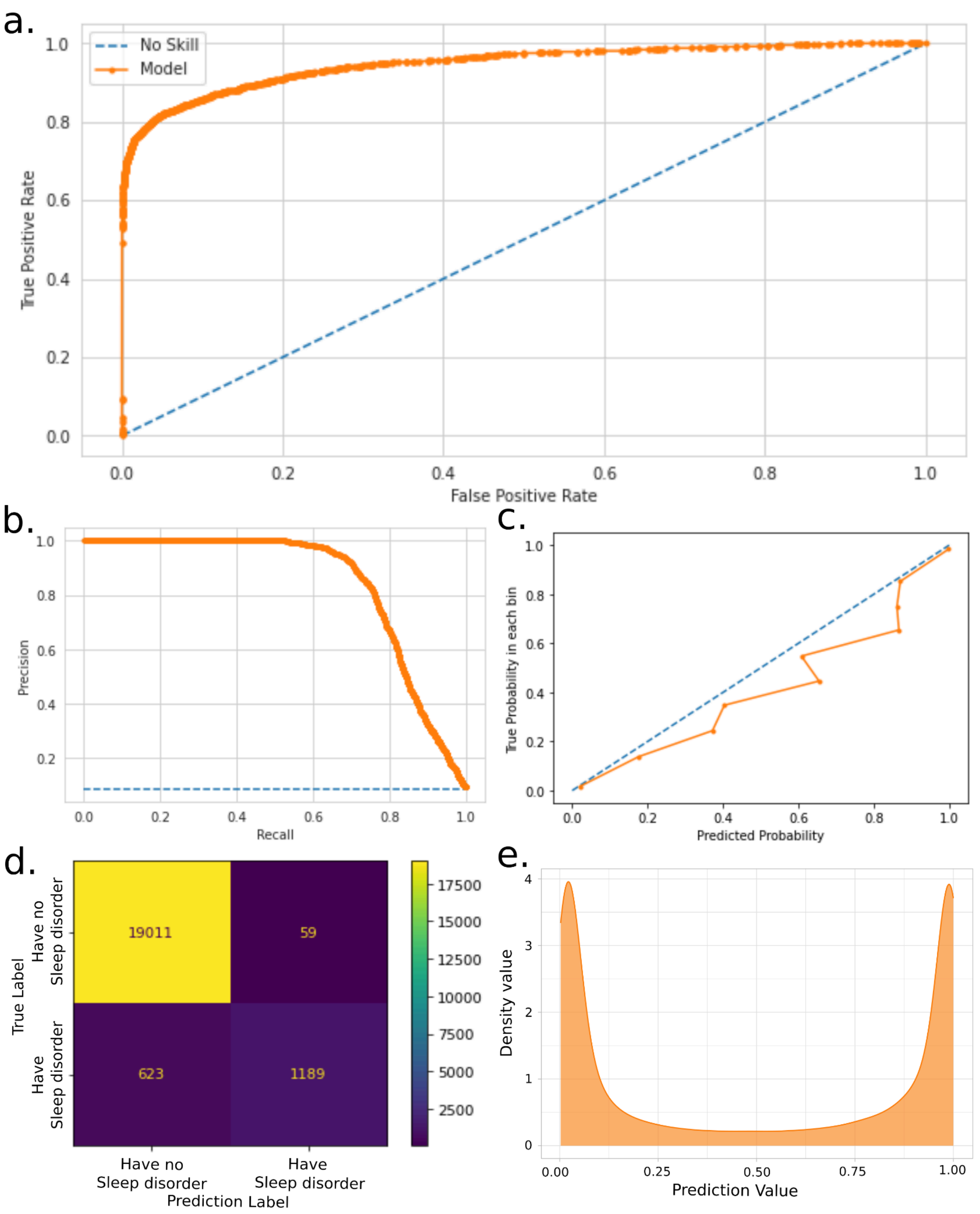


**Supplementary Fig. 14 | Evaluation of XGBoost model performance in predicting Sleep disorder.**

Performance metrics for XGBoost classifier trained to distinguish sleep disorder cases from individuals with no recorded mental health diagnoses (super control definition). Model evaluation was conducted on a held-out validation set comprising 20% of the data. a) Receiver operating characteristic (ROC) curve (AUC = 96.73%). b) Precision-Recall curve. c) Calibration curve for predicted probabilities and observed outcomes. d) Confusion matrix displaying classification results on the validation set. e) Density plot showing predicted probabilities for those excluded from the super control phenotype.

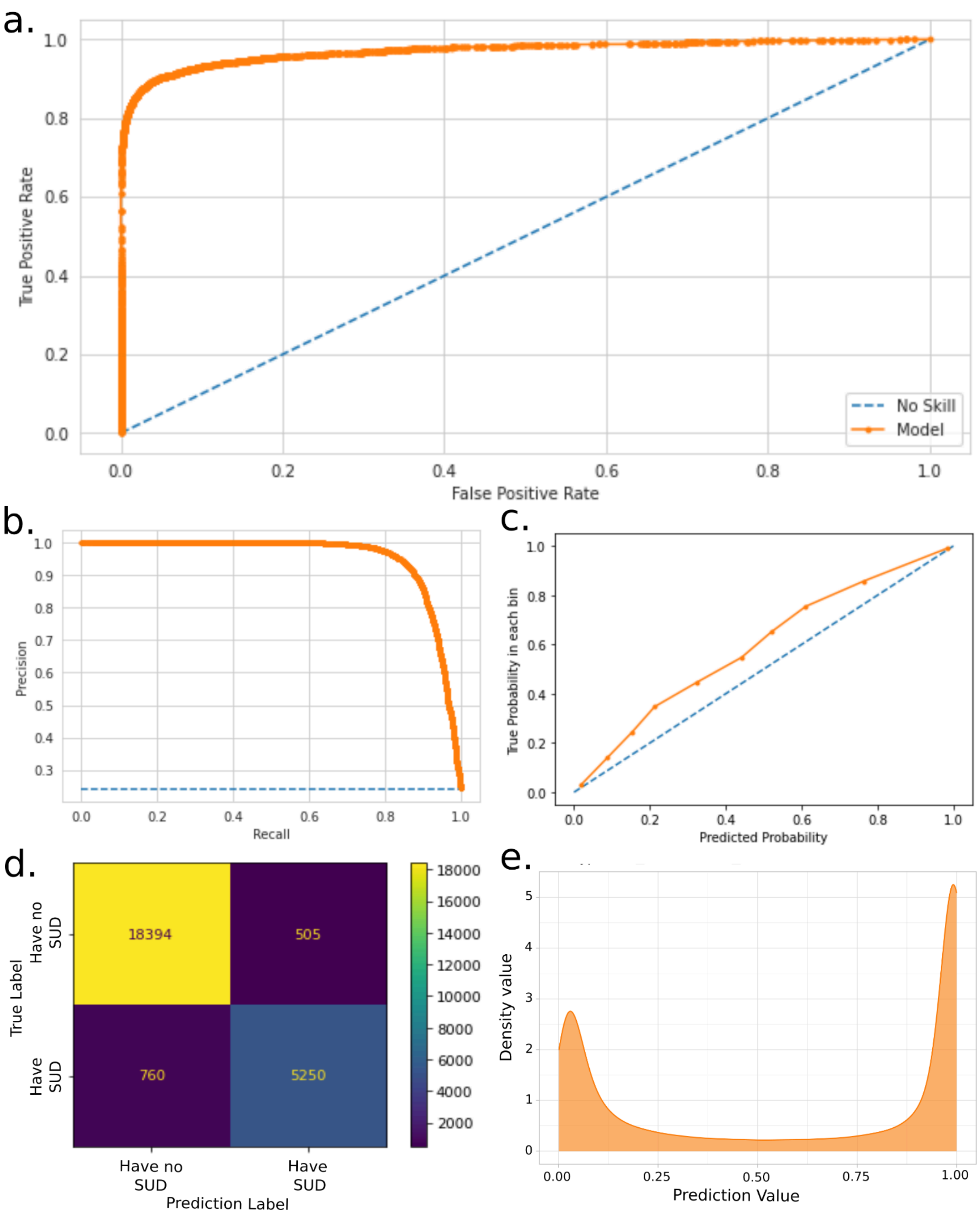


**Supplementary Fig. 15 | Evaluation of XGBoost model performance in predicting SUD.**

Performance metrics for XGBoost classifier trained to distinguish SUD cases from individuals with no recorded mental health diagnoses (super control definition). Model evaluation was conducted on a held-out validation set comprising 20% of the data. a) Receiver operating characteristic (ROC) curve (AUC = 94.92%). b) Precision-Recall curve. c) Calibration curve for predicted probabilities and observed outcomes. d) Confusion matrix displaying classification results on the validation set. e) Density plot showing predicted probabilities for those excluded from the super control phenotype.

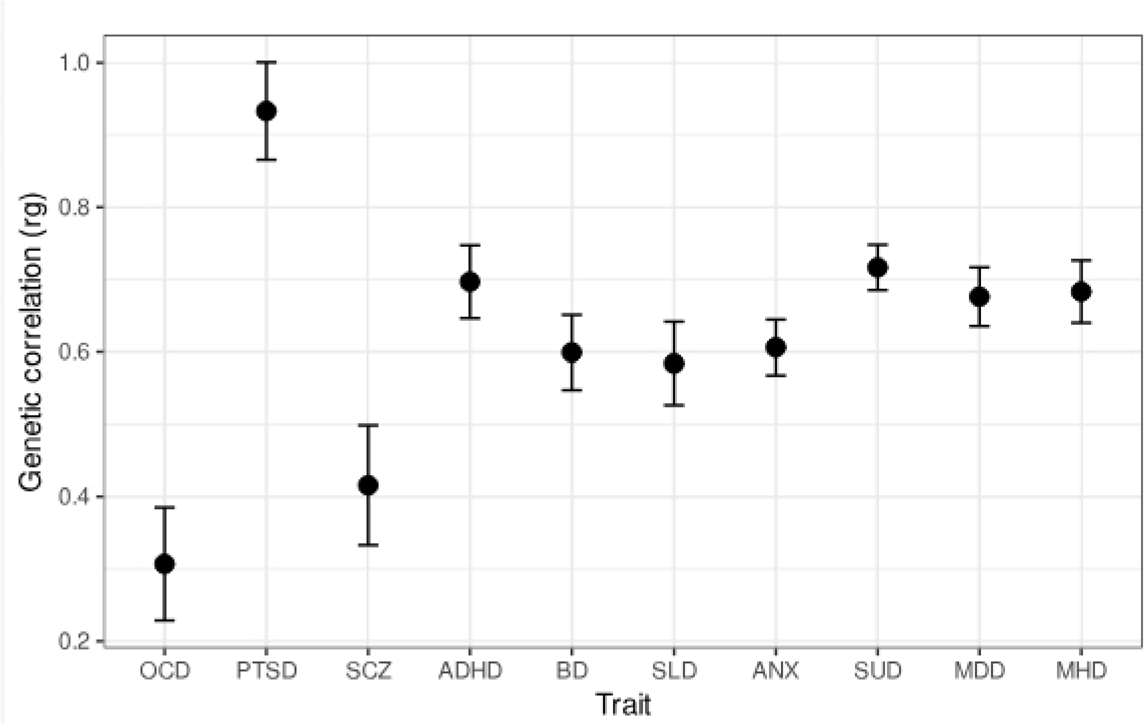


**Supplementary Fig. 16 | Genetic correlations across psychiatric traits between super control and predicted excluded cohort.**

Genetic correlations (rg) were estimated using LDSC between GWAS results from super control phenotype and the predicted excluded cohort for the ten traits with standard error displayed as error bars.

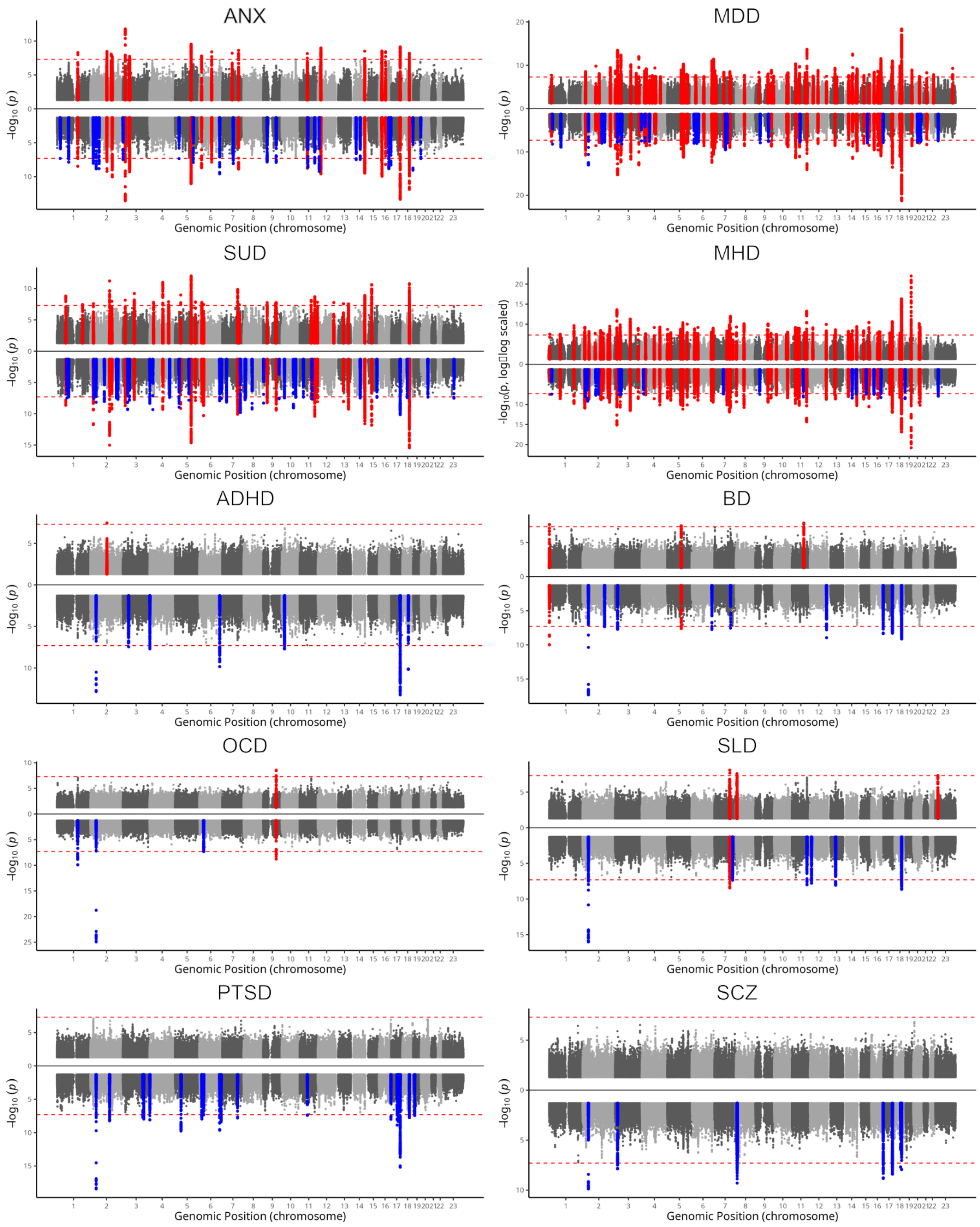


**Supplementary Fig. 17 | Manhattan plot of Genome-wide significant loci in Original FinnGen and SuperControl phenotypes across psychiatric traits.**

a-f, Mirrored Manhattan plots for anxiety (ANX), major depressive disorder (MDD), substance use disorder (SUD), a combined “any mental health disorder” trait (MHD), attention-deficit/hyperactivity disorder (ADHD), bipolar disorder (BD), obsessive-compulsive disorder (OCD), sleep disorders (SLD), post-traumatic stress disorder (PTSD) and schizophrenia (SCZ). In each plot, the upper half represents results from the SuperControl phenotype and the lower half the PRISMA phenotype. Red points denote loci shared between the two phenotypes or unique to the SuperControl phenotype, and blue points denote loci unique to the PRISMA phenotype.

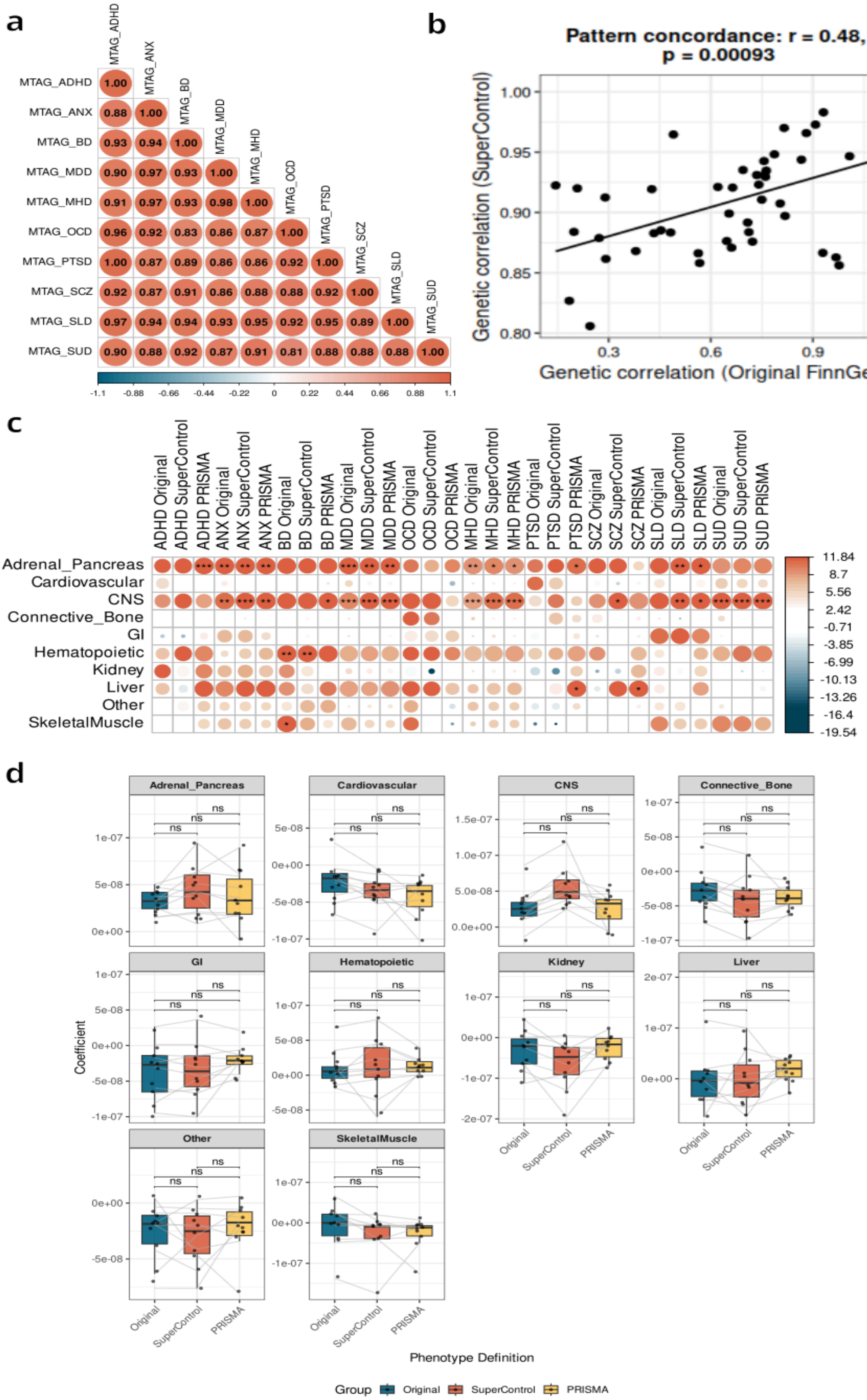

a
MTAG_ADHD
MTAG_ANX
MTAG_BD
MTAG_MDD
MTAG_MHD
MTAG_OCD
MTAG_PTSD
MTAG_SCZ
MTAG_SLD
MTAG_SUD
1.00
0.88 1.00
0.93 0.94 1.00
0.90 0.97 0.93 1.00
0.91 0.97 0.93 0.98 1.00
0.96 0.92 0.83 0.86 0.87 1.00
1.00 0.87 0.89 0.86 0.86 0.92 1.00
0.92 0.87 0.91 0.86 0.88 0.88 0.92 1.00
0.97 0.94 0.94 0.93 0.95 0.92 0.95 0.89 1.00
0.90 0.88 0.92 0.87 0.91 0.81 0.88 0.88 0.88 1.00
-1.1 -0.88 -0.66 -0.44 -0.22 0 0.22 0.44 0.66 0.88 1.1
b
Pattern concordance: r = 0.48, p = 0.00093
Genetic correlation (SuperControl)
Genetic correlation (Original FinnGen)
c
ADHD Original
ADHD SuperControl
ADHD PRISMA
ANX Original
ANX SuperControl
ANX PRISMA
BD Original
BD SuperControl
BD PRISMA
MDD Original
MDD SuperControl
MDD PRISMA
OCD Original
OCD SuperControl
OCD PRISMA
MHD Original
MHD SuperControl
MHD PRISMA
PTSD Original
PTSD SuperControl
PTSD PRISMA
SCZ Original
SCZ SuperControl
SCZ PRISMA
SLD Original
SLD SuperControl
SLD PRISMA
SUD Original
SUD SuperControl
SUD PRISMA
Adrenal_Pancreas
Cardiovascular
CNS
Connective_Bone
GI
Hematopoietic
Kidney
Liver
Other
SkeletalMuscle
11.84 8.7 5.56 2.42 -0.71 -3.85 -6.99 -10.13 -13.26 -16.4 -19.54
d
Adrenal_Pancreas
Cardiovascular
CNS
Connective_Bone
GI
Hematopoietic
Kidney
Liver
Other
SkeletalMuscle
ns
Coefficient
Original
SuperControl
PRISMA
Phenotype Definition
Group Original SuperControl PRISMA

**Fig. 18 | Genetic correlations and tissue-specific heritability across refined phenotypes.**
All panels show results for ten psychiatric traits: obsessive–compulsive disorder (OCD), post-traumatic stress disorder (PTSD), attention-deficit/hyperactivity disorder (ADHD), schizophrenia (SCZ), major depressive disorder (MDD), anxiety (ANX), substance use disorder (SUD), sleep disorder (SLD), bipolar disorder (BD), and the combined "any mental health disorder" trait (MHD).
(a) Heatmap of pairwise genetic correlations among traits for the PRISMA phenotype.
(b) Scatter plot comparing pairwise genetic correlations across ten psychiatric traits estimated using LDSC for the Original FinnGen and SuperControl definitions. Each point represents one trait pair. The overall correlation between definitions was moderate but significant (Pearson $r = 0.48$, $p = 9.3 \times 10^{-4}$).
(c) Heatmap of partitioned heritability enrichment across cell types using the baseline cell-type dataset. Columns represent traits, and rows represent cell types; asterisks indicate significant enrichments.
(d) Boxplot of partitioned heritability coefficients across major tissue groups. Colors indicate phenotype definitions: Original FinnGen (blue), SuperControl (red), and PRISMA (yellow). Significance was assessed using paired Wilcoxon rank-sum tests comparing effect sizes between definitions.

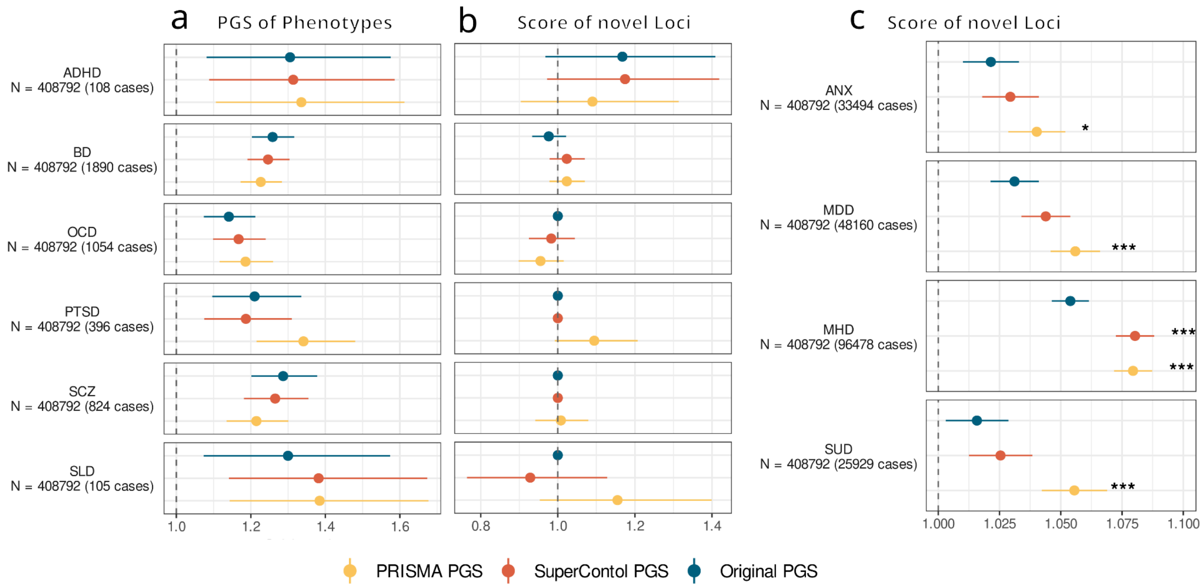


**Fig. 19 | Polygenic score prediction in the UK Biobank.**
a, Odds ratios (with standard errors) from polygenic score prediction for 6 psychiatric traits, SCZ, BD, ADHD, PTSD, OCD and SLD.
b, Odds ratios (with standard errors) for the same 6 traits when restricting to SNPs not replicated in the locus-based replication (LBR) of the respective phenotypes.
c, Odds ratios (with standard errors) for 4 psychiatric traits, ANX, MDD, MHD, and SUD when restricting to SNPs not replicated in the locus-based replication (LBR) of the respective phenotypes.
All results are shown for the SuperControl (red), Original FinnGen (blue), and PRISMA (yellow) GWAS
